# Multimodal Machine Learning–Based Clinical and Genomic Prognostic Factors for Progression-Free Survival in Metastatic Breast Cancer Using Phase III MONALEESA Trials

**DOI:** 10.64898/2026.10.09.26364892

**Authors:** Craig Wang, Rahul Paul, Yutong Li, Sean Smith, Qian Cao, Stephen H. Chang, Rosane Charlab, Thibaud Coroller, Ruihao Huang, Laleh Amiri-Kordestani, Zheng Li, Qi Liu, Christy Osgood, Severine Peyrard, Berkman Sahiner, Luis Santana-Quintero, Faye Su, Juan P. Zarate, Hao Zhu, David Ohlssen, Nicholas Petrick, Anup Amatya, Nadia Solovieff

**Author notes:** **Corresponding authors:** Nadia Solovieff, Advanced Quantitative Sciences, Novartis Pharmaceutical Company, 250 Massachusetts Avenue, Cambridge, Massachusetts 02139, United States.; Rahul Paul, Office of Biostatistics and Pharmacovigilance, Center for Biologics Evaluation and Research, Food and Drug Administration, 10903 New Hampshire Avenue, Silver Spring, Maryland 20993, United States. Joint first author (Craig Wang, Rahul Paul). Joint second author (Yutong Li, Sean Smith). Joint senior author (Anup Amatya, Nadia Solovieff). **Disclaimer**: This article reflects the views of the authors and does not represent the views or policy of the US Food and Drug Administration, the Department of Health and Human Services, or the US Government. The mention of commercial products, their sources, or their use in connection with material reported herein is not to be construed as either an actual or implied endorsement of such products by the Department of Health and Human Services. **Conflict of interest:** All disclosures will be added to the final publication of the manuscript in a peer-reviewed journal.

## Abstract

**Purpose:** Cyclin-dependent kinase 4/6 inhibitors in combination with endocrine therapy have become widely adopted as a first-line treatment approach for hormone receptor-positive (HR+)/human epidermal growth factor receptor 2-negative (HER2−) advanced breast cancer (ABC). Through a collaboration between Novartis and the US Food and Drug Administration, we integrated multimodal data from phase III MONALEESA trials to identify novel prognostic factors for HR+/HER2−ABC.

**Experimental Design:** Baseline clinical and genomics data were collected from patients in the MONALEESA-2, −3, and −7 trials. Circulating tumor DNA (ctDNA) from plasma samples and gene expression in archival tumor biopsies were analyzed. Machine learning models were trained (*n* = 1,355) on progression-free survival (PFS) using multimodal features via fusion models. In a test set (*n* = 382), prognostic performances were evaluated using the Kaplan-Meier, Cox, and Weibull models.

**Results:** The late fusion model achieved a prognostic performance (C-index: 0.727, 95% CI: 0.641–0.799) slightly outperforming early fusion (C-index: 0.718, 95% CI: 0.631–0.791) and single-modality models. Model-based risk stratification identified distinct low- and high-risk groups (PFS HR, 0.28; 95% CI: 0.20–0.38). Among the top 40 features for progression risk (18 clinical, 20 gene expression, and 2 ctDNA), these were associated with higher risk: liver involvement, higher histological grades, early relapse; lower *STC2 and RBBP8* expressions and higher *FBX05* expression; and higher tumor mutational burden and ctDNA purity.

**Conclusions:** Integrating clinical and genomic modalities improved progression risk prediction compared with single-modality models, while highlighting salient features that inform the underlying biology and support hypothesis generation.

## Introduction

Breast cancer is the second most frequent cause of cancer death in women (1). In 2026, it was estimated that 324,580 new cases of invasive breast cancer were expected to be diagnosed in the United States, with approximately 42,670 deaths expected (2). Hormone receptor-positive (HR+), human epidermal growth factor receptor 2-negative (HER2−) breast cancer is the most common subtype of breast cancer accounting for ∼70% of cases (3). While endocrine therapy (ET) remains the backbone for the treatment of HR+/HER2− advanced breast cancer (ABC), NCCN Clinical Practice Guidelines in Oncology (NCCN guidelines^®^) included cyclin-dependent kinases 4 and 6 (CDK 4/6) inhibitors, which target the cell cycle and prevent cancer cell division, in combination with an aromatase inhibitor or fulvestrant as first-line options (4).

Ribociclib, a CDK 4/6 inhibitor, has been investigated across a broad population of patients with HR+/HER2− ABC, covering pre- and post-menopausal women, first- or second-line therapy, and different backbone ET including aromatase inhibitors, fulvestrant, and tamoxifen in the MONALEESA-2, MONALEESA-3, and MONALEESA-7 randomized, double-blind, placebo-controlled, international phase III clinical trials (5–7). Based on the results of these trials, ribociclib was approved for the treatment of HR+/HER2− ABC as initial endocrine-based therapy or following disease progression on ET (8, 9).

Prognostic factors that can accurately predict outcomes of patients have an important impact on both treatment decisions by clinicians and personal decisions by patients. There are several well-established clinical prognostic factors in HR+/HER2− ABC, including progesterone receptor status, higher tumor grade, higher circulating tumor count and Ki67 level, number of metastatic sites, presence of visceral metastases, and prior treatment with ET, that are commonly used as stratification factors in clinical trials (10, 11). Although factors such as these have historically been the foundation of prognostic assessment in patients with cancer, recent advances in statistical learning and artificial intelligence have enabled the integration of complex, multimodal data to improve outcome prediction across cancer types. Models combining clinical variables with genomic and multiomics data have demonstrated improved prognostic and predictive results in several malignancies, including urothelial carcinoma, colon cancer, and glioblastoma (12–14). Additionally, deep learning models that leverage multimodal data have outperformed conventional risk stratification tools and identified novel prognostic features in prostate, colorectal, cervical, and hepatocellular cancers (15–18). Collectively, these studies highlight the growing role of machine learning–based models in advancing individualized prognostic assessment and treatment decision making in oncology. Nonetheless, identification of additional and improved prognostic factors may further improve clinical utility of the models.

Statistical and machine learning approaches that integrate complex, multimodal data sets have enabled the identification of novel prognostic factors across multiple cancer types. For example, analyses combining clinical and genomic variables have demonstrated utility in predicting response to immune checkpoint inhibition in advanced urothelial carcinoma (13). Similarly, integration of clinical data with multiomics profiles has been shown to improve prognostic results in colon cancer, with measurable gains in C-index compared with clinical data alone, and to enhance overall survival (OS) prediction in patients with isocitrate dehydrogenase–wildtype glioblastoma (12, 14). Advances in deep learning have further expanded prognostic modeling capabilities. Self-supervised convolutional neural networks trained on clinical and histopathological data have outperformed established guideline-based risk assessment tools in predicting prostate cancer development (16). In metastatic colorectal cancer, artificial intelligence (AI)–augmented deep learning models have successfully stratified patients receiving first-line therapy into distinct risk groups and identified carcinoembryonic antigen, neutrophil-to-lymphocyte ratio, and liver function parameters as key progression-free survival (PFS) predictors (18). Large population-based data sets have also been leveraged to develop machine learning models with strong prognostic performance; models trained on Surveillance, Epidemiology, and End Results (SEER) registry data achieved Areas Under the Receiver Operating Characteristic curve (AUCs) of >0.9 in predicting 1-, 3-, and 5-year OS in cervical cancer and were proposed as tools to support individualized treatment planning (17). More recently, AI-assisted Cox regression approaches have been applied to identify prognostic factors in patients with hepatocellular carcinoma treated with combination immunotherapy and targeted therapies, with or without locoregional interventions (15).

In 2020, Novartis Pharmaceuticals Corporation and the US Food and Drug Administration (FDA) began a multiyear scientific collaboration under a Research Collaboration Agreement to use the MONALEESA series of clinical trials to find novel prognostic and predictive factors for HR+/HER2− ABC (19). In this manuscript, we employ and compare the modeling results from various analytical approaches, including dimensionality reduction, regression, random forest, deep learning, and model fusion, to identify factors associated with PFS, the primary endpoint of MONALEESA-2, −3, and −7.

Building on this growing body of evidence, our study extends the application of multimodal prognostic modeling to a phase III clinical trial population, highlighting the potential for such approaches to inform risk stratification and trial design in HR+/HER2− ABC.

## Methods

### Patients and studies

Patients were enrolled between 2013 and 2016 across MONALEESA-2 (NCT01958021), MONALEESA-3 (NCT02422615), and MONALEESA-7 (NCT02278120) trials; all patients were included in the analyses. Key baseline demographics and disease characteristics of the three studies are presented in (20). All three MONALEESA trials evaluated ribociclib in patients with HR+/HER2− advanced or metastatic breast cancer. MONALEESA-2 enrolled postmenopausal women with no prior ET or chemotherapy for ABC, and patients received either ribociclib or placebo in combination with letrozole; median follow-up for the final PFS analysis was 15.3 months (21). MONALEESA-3 enrolled postmenopausal patients with ≤1 prior line of ET and no prior chemotherapy for ABC, and patients received fulvestrant plus either ribociclib or placebo; median follow-up for the final PFS analysis was 20.4 months (22). MONALEESA-7 recruited premenopausal or perimenopausal women with no prior ET for ABC (≤1 prior line of chemotherapy was permitted), and patients received either ribociclib or placebo combined with ET (either a nonsteroidal aromatase inhibitor or tamoxifen ± goserelin); median follow-up for the final PFS analysis was 19.2 months (23). Analyses used data collected up to the final PFS analysis of each study. Data were anonymized prior to analysis (24).

### Data collection

Baseline clinical data such as demographics, cancer history, and laboratory results were collected using an electronic case report form. Baseline genomics data including both DNA sequencing of circulating tumor DNA (ctDNA) and gene expression data from archival tumor biopsies were analyzed (details in the Supplementary Material).

In all three studies, tumor assessments were performed every 8 weeks for the first 18 months and every 12 weeks subsequently. The outcome of interest, PFS was defined as the time from randomization to disease progression or death due to any cause and was assessed by the local investigator per Response Evaluation Criteria in Solid Tumors (RECIST) v1.1. Patients without disease progression or death as of the final PFS analyses were censored at the date of the last tumor assessment.

### Feature engineering

Clinical data included baseline demographics, prior therapy, extent of cancer, and laboratory assessments were pre-processed following Clinical Data Interchange Standards Consortium standards during clinical trial reporting activities. Features with duplicated information (e.g., age and age group) and with >10% missingness were not considered. The final clinical data set contained 72 baseline clinical features, including 28 categorical and 44 continuous features.

The analysis of ctDNA data focused on single base mutations, short indels, and copy alterations. Since different types of variants can have different functional impact on the downstream protein, data were collapsed at the gene level to create 3 features per gene: amplified, truncating (nonsense or frameshift), or other (missense, splice, or non-frameshift). These were coded as 1 if present and 0 if not. We excluded ctDNA features with a frequency <3% from the modeling. There were 32 features remaining for modeling training after the above preprocessing step. Tumor mutation burden and purity, an estimate of the amount of tumor DNA in the circulation, were also included.

Normalized gene expression values were included for each gene (781 features), along with an additional feature indicating whether the patient received an intervening line of therapy between the biopsy and trial start, given the variability in the timing of tumor biopsy collection (see Supplementary Material for details).

### Analytical methods

#### Data split

Data were partitioned into training set (sample size depends on modality) used for model fitting and tuning, evaluation set (*n* = 242) used during model development for performance comparison, and testing set (*n* = 382) used as an independent heldout data set only for the final performance assessment and interpretation. The split used stratified random sampling by data availability and study to ensure representation across trials and data modalities (**Supplementary Table S1**) (25).

#### Initial feature selection

Univariate Cox regression was used as a variable selection step within a stability selection approach to identify robust prognostic features (26). For each modality (ctDNA, gene expression, and clinical), 50 randomly generated stratified 70/30 splits of the training set (based on the PFS event indicator) were created. Feature selection was conducted exclusively on the 70% subset in each iteration, with the remaining 30% withheld to mitigate overfitting. Following the stability selection step (26), features were considered stable if they were selected in more than 70% of the 50 resampled subsets with a *P*-value < 0.05. Stable feature selection was applied to the elastic net and survival random forest models, but not to the deep learning model, in order to allow it to capture complex nonlinear and latent feature interactions. As a sensitivity analysis, a more stringent *P*-value threshold of < 0.01 was also evaluated.

#### Individual modality model training

For each modality, three different classes of the model were evaluated: regression (i.e., elastic net), tree-based (i.e., survival random forest), and deep learning (27, 28) (i.e., multilayer perceptron (29)). Cross-validation was repeated 50 times for each combination of tuning parameters. For each model, the respective hyperparameters were tuned based on the respective loss functions, that is, negative partial log-likelihood for elastic net and multilayer perceptron, and log rank test statistic for survival random forest. Harrell’s C-index, which evaluates the discrimination performance of models by assigning a higher risk to the patient that experiences an event first from any random pair of patients, was used for model evaluation (30). The best performing model in terms of concordance-index (C-index) were reported.

Using the selected hyperparameters, the model was trained on the full training set and evaluated on the evaluation set. The top-performing individual-modality model was selected based on its C-index on the evaluation set. The selected model was subsequently re-trained on all available training data (training and evaluation sets combined, up to *n* = 1,355 depending on modality, see **Supplementary Table S1**) to increase the sample size. This retraining step did not involve additional feature selection or re-tuning of hyperparameters. The retrained model was then used for final independent testing on held out testing set.

#### Multimodality (fusion) model training

Early fusion was used to combine the data modalities into a single model, where all the variables from each modality were considered jointly in the feature selection process (following the same cross validation process as individual modality training). Early fusion allows the model to learn joint interactions across modalities but is limited to patients with complete multimodal data (*n =* 336). As an alternative to early fusion, we also applied a late fusion model, which used standardized predictions from top-performing modality-specific models as input features and assigned modality-specific weights through Weibull regression. The late fusion model acted as an ensemble of three individual models and preserves sample size by combining modality-specific risk predictions while retaining interpretability of each data source.

An overall workflow of the multimodal analytical pipeline is illustrated (**Fig. 1**).

**Figure 1.**
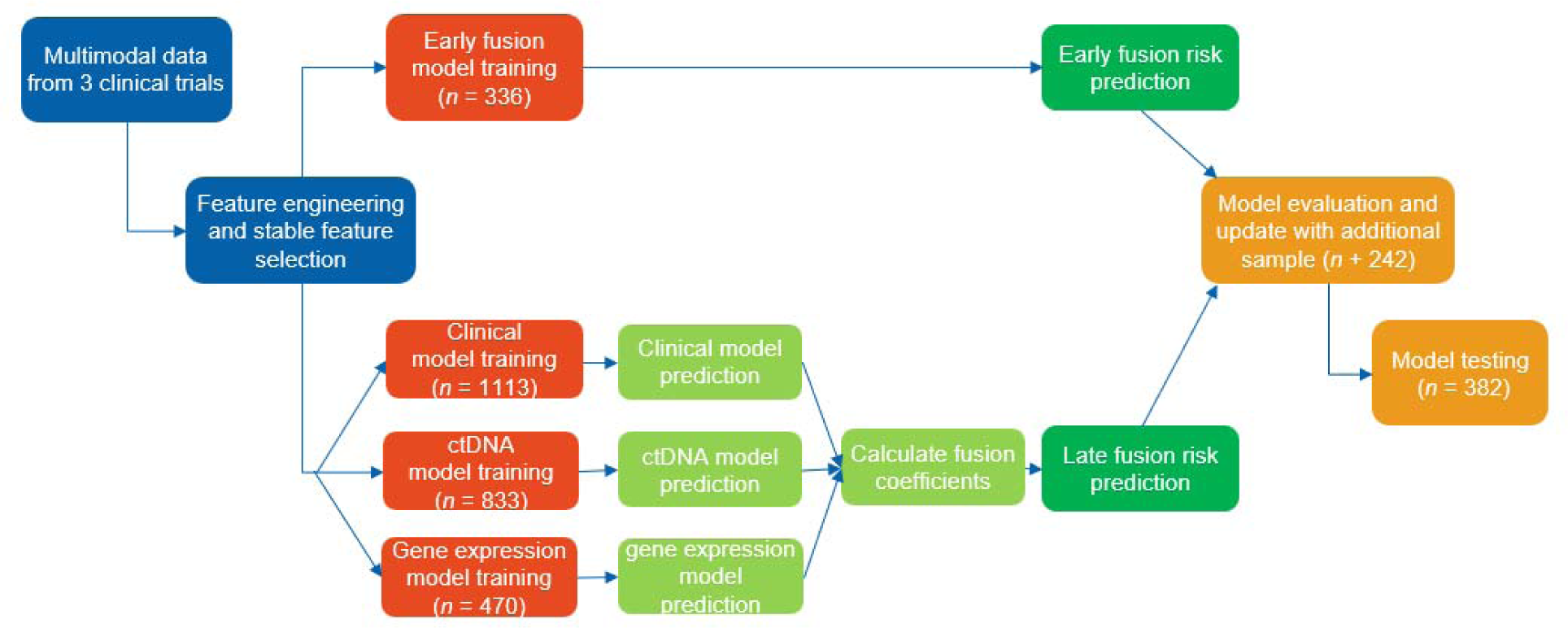
Illustration of the multimodal fusion analytical pipeline. ctDNA, circulating tumor DNA.

### Interpretation methods

The selected best performing model was applied to the test set, and each patient was classified into different risk groups (low vs. high) based on median predicted risk score. PFS probability was estimated using the Kaplan-Meier method to compare the risk groups overall, as well as separately for each treatment arm for isolated prognostic effect evaluation since treatment arm was included as a feature during model training. Forest plot with hazard ratios were computed using Cox proportional hazard model.

Another approach to evaluate the performance of predicted risk score is through covariate adjusted treatment effect estimation (31). The predicted risk scores were incorporated as covariates in the analysis of PFS in the test set. As an intermediate step, the original risk scores were transformed by taking residuals from a linear model with treatment arm as a single covariate to remove treatment arm information that was used during multimodal training. These transformed risk scores were then included as a covariate in a Cox proportional hazards model to adjust for baseline prognostic differences among patients. The adjusted analysis was compared to an unadjusted Cox model to quantify gains in terms of % effective sample size, approximated using percentage variance reduction in log (hazard ratio [HR]) estimates.

Additional interpretable machine learning methods such as Shapley additive explanation (SHAP) score (32) were also used to quantify the additive contribution of individual baseline covariates from clinical and biomarker data to the predicted risk score. The late fusion model utilized a Weibull regression as the last step, which predicts the probability of events at any given timepoint for each patient. Given the duration of follow-up in the trials, 12 months was chosen as the landmark time, and the prediction was compared with the observed proportion of events at four different quartiles to assess accuracy.

These analyses were conducted using Python and R. Further details of the methods are provided in the Supplementary Methods.

## Results

### Model development and selection during training

When testing the candidate models, the early and late fusion models performed consistently better than any of the models based on individual data modalities (**Table 1**). During training, the best performing models were deep learning early fusion model (cross validated C-index, 0.708; C-index on evaluation set, 0.749) and late fusion model (C-index on evaluation set, 0.746). The late fusion results indicate that each modality contributes independently to the prediction (*P* < 0.001 for each modality), and the coefficients quantify each modality’s contribution to the final prediction (**Table 1**), with the strongest effect for gene expression and the weakest for ctDNA. Based on these results, the two top performing models, namely the deep learning early fusion model and Weibull-weighted late fusion model (with deep learning model for clinical and elastic net for ctDNA and gene expression) models were selected for further testing.

**Table 1.** Model performance during development and independent test evaluation.

|  | Model performance measured by C-index |  |  |
| --- | --- | --- | --- |
| Model class | Cross-validated training set (mean, SD) | Evaluation set (N = 242) | Test set (95% CI) (N = 382) |
| **Individual modality** |  |  |  |
| **Clinical** |  |  |  |
| Elastic net | 0.677 (0.021) | 0.664 | - |
| Random forest | 0.682 (0.020) | 0.646 | - |
| Deep learning | 0.669 (0.019) | 0.699 | - |
| **ctDNA** |  |  |  |
| Elastic net | 0.669 (0.018) | 0.652 | - |
| Random forest | 0.658 (0.019) | 0.648 | - |
| Deep learning | 0.659 (0.016) | 0.656 | - |
| **Gene expression** |  |  |  |
| Elastic net | 0.693 (0.031) | 0.707 | - |
| Random forest | 0.684 (0.034) | 0.640 | - |
| Deep learning | 0.674 (0.022) | 0.674 | - |
| **Multimodal fusion** |  |  |  |
| **Early fusion** |  |  |  |
| Regression | 0.777 (0.027) | 0.716 | - |
| Random forest | 0.765 (0.035) | 0.694 | - |
| Deep learning | 0.708 (0.035) | 0.749 | 0.718 (0.631 –0.791) |
| **Late fusion<sup>a</sup>** | NA | 0.746 | 0.727 (0.641 –0.799) |

<sup>a</sup>Late fusion was based on clinical deep learning model, ctDNA elastic net model and gene expression elastic net model using Weibull regression weights. C-index, concordance index; ctDNA, circulating tumor DNA; NA, not applicable; SD, standard deviation.

### Testing and risk stratification

The selected models were retrained with the combined training and evaluation data sets used are part of algorithm development. Final model performance was assessed using the independent test data set, which was not used during any stage of model development. The C-index in the independent test set was 0.718 (95% CI: 0.631–0.791) for the early fusion deep learning model and 0.727 (95% CI: 0.641–0.799) for the late fusion model (**Table 1**). Given the slightly better C-index and especially the easier interpretation of the late fusion model, it was chosen as the final model for interpretation.

Using the late fusion models, a predicted risk score for each patient in the test set was computed and used to classify patients as low- or high-risk by the median risk in the training set. The HR for PFS in low-vs high-risk patients were 0.28 (95% CI, 0.20–0.38), and the observed event rate was 52/198 (26.3%) versus 122/184 (66.3%), respectively (**Fig. 2**). Across known prognostic subgroups, the prognostic value of the predicted score from the model remained strong (**Supplementary Fig. S1**).

**Figure 2.**
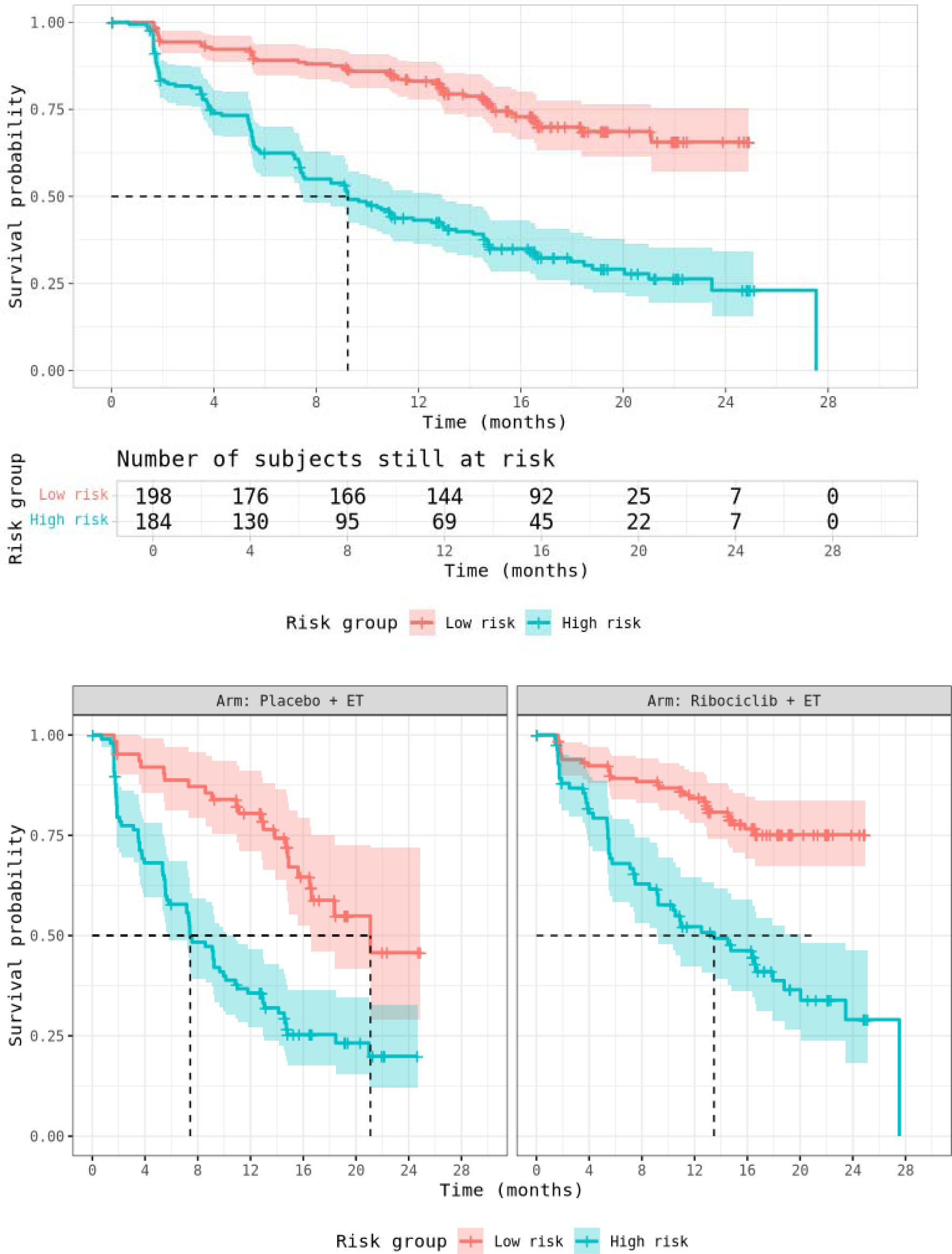
Kaplan-Meier curves of progression-free survival by model-defined risk group in the test data set, overall and by treatment arms. ET, endocrine therapy.

As an alternative method to evaluate the prognostic value, incorporating the transformed risk scores as a covariate adjustment when estimating treatment effect resulted in notable efficiency gains consistent with results based on C-index. Specifically, the adjusted analysis yielded a 17% increase in effective sample size compared to the unadjusted Cox model. However, while we used transformed risk scores as an approach for model evaluation, the validity of these scores for prospective use remains to be determined.

### Model interpretation

Overall, the late fusion weights suggest that gene expression contributed most strongly to the combined prognostic signal, followed by clinical features, with ctDNA providing a smaller but still significant contribution (**Table 2**). Among the top 40 most important features in the late fusion model, 18 were clinical, 20 were gene expression related, and 2 were ctDNA related. The randomized treatment arm assignment was identified as the most important feature (higher PFS risk with placebo; **Fig. 3**). The SHAP plots of individual models contributing to the late fusion are presented in **Supplementary Fig. S2**. The most important clinical features were liver involvement (higher risk associated with presence), histological grade (higher risk associated with higher grade), and prior ET (higher risk of progression within 12 months of prior ET). The most important gene expression–related features were *STC2* (higher expression associated with better PFS), *RBBP8* (higher expression associated with better PFS), and *FBXO5* (higher expression associated with worse PFS). In the gene expression model, the presence of an intervening line of therapy between tumor biopsy and trial treatment initiation (N_therapy) was also identified as an important feature. The most important ctDNA features were tumor mutational burden (TMB; higher TMB associated with worse PFS) and purity (higher purity associated with worse PFS).

**Figure 3.**
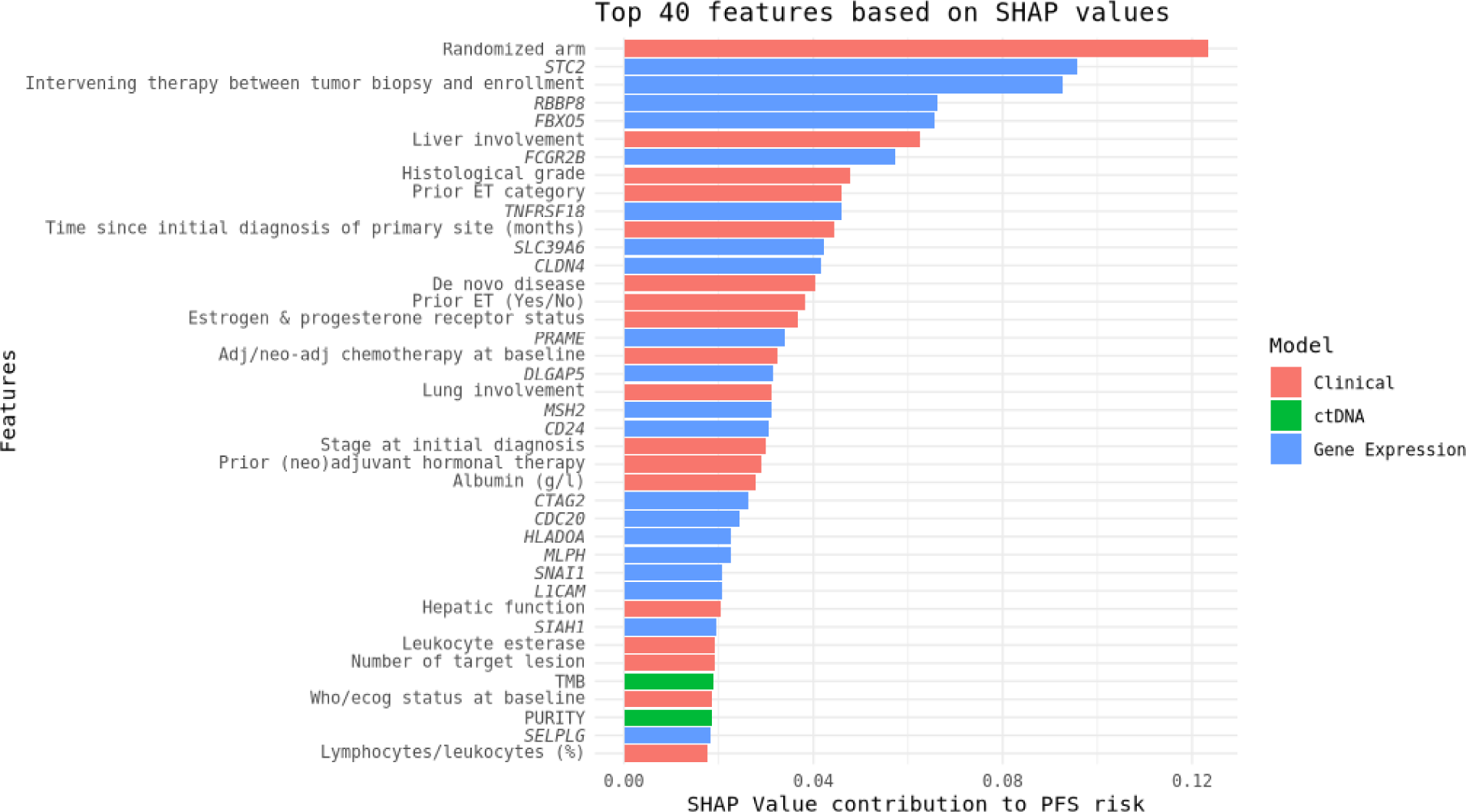
Top 40 features (SHAP)—late fusion. adj, adjuvant; CD24, cluster of differentiation 24; CDC20, cell division cycle 20; CLDN4, claudin 4; CTAG2, cancer/testis antigen 2; ctDNA, circulating tumor DNA; DLGAP5, discs large–associated protein 5; ET, endocrine therapy; FBXO5, Fbox protein 5; FCGR2B, Fc gamma receptor IIB; HLADOA, HLA class II histocompatibility antigen DO alpha chain; L1CAM, L1 cell adhesion molecule; MLPH, melanophilin; MSH2, MutS homolog 2; PRAME, preferentially expressed antigen in melanoma; Prior ET category, prior endocrine therapy category; PURITY, ctDNA purity; RBBP8, retinoblastoma binding protein 8; SELPLG, Selectin P ligand; SHAP, shapely additive explanation; SIAH1, SIAH e3 ubiquitin protein ligase 1; SLC39A6, solute carrier family 39 member 6; SNAI1, snail family transcriptional repressor 1; STC2, stanniocalcin 2; TMB, tumor mutational burden; TNFRSF18, tumor necrosis factor receptor superfamily member 18; WHO/ECOG, World Health Organization/Eastern Cooperative Oncology Group.

**Table 2.** Late fusion model weights of standardized risk scores derived from individual data modalities.

| **Data modality** | **Weights** | **95% CI<sup>a</sup>** | ***P*-value** |
| --- | --- | --- | --- |
| Clinical | 0.44 | 0.34–0.55 | <0.001 |
| ctDNA | 0.18 | 0.11–0.26 | <0.001 |
| Gene expression | 0.50 | 0.40–0.60 | <0.001 |

<sup>a</sup> The weights are based on the Weibull regression coefficients on the shape parameter. ctDNA, circulating tumor DNA.

Given that prediction of PFS probability is a more clinically meaningful metric than ranked risk predictions, the prediction based on the late fusion Weibull regression model was converted into a probability of PFS at 12 months. Patients categorized as having higher model-predicted probabilities of PFS at 1 year showed correspondingly higher observed rates of 12-month PFS (**Fig. 4**).

**Figure 4.**
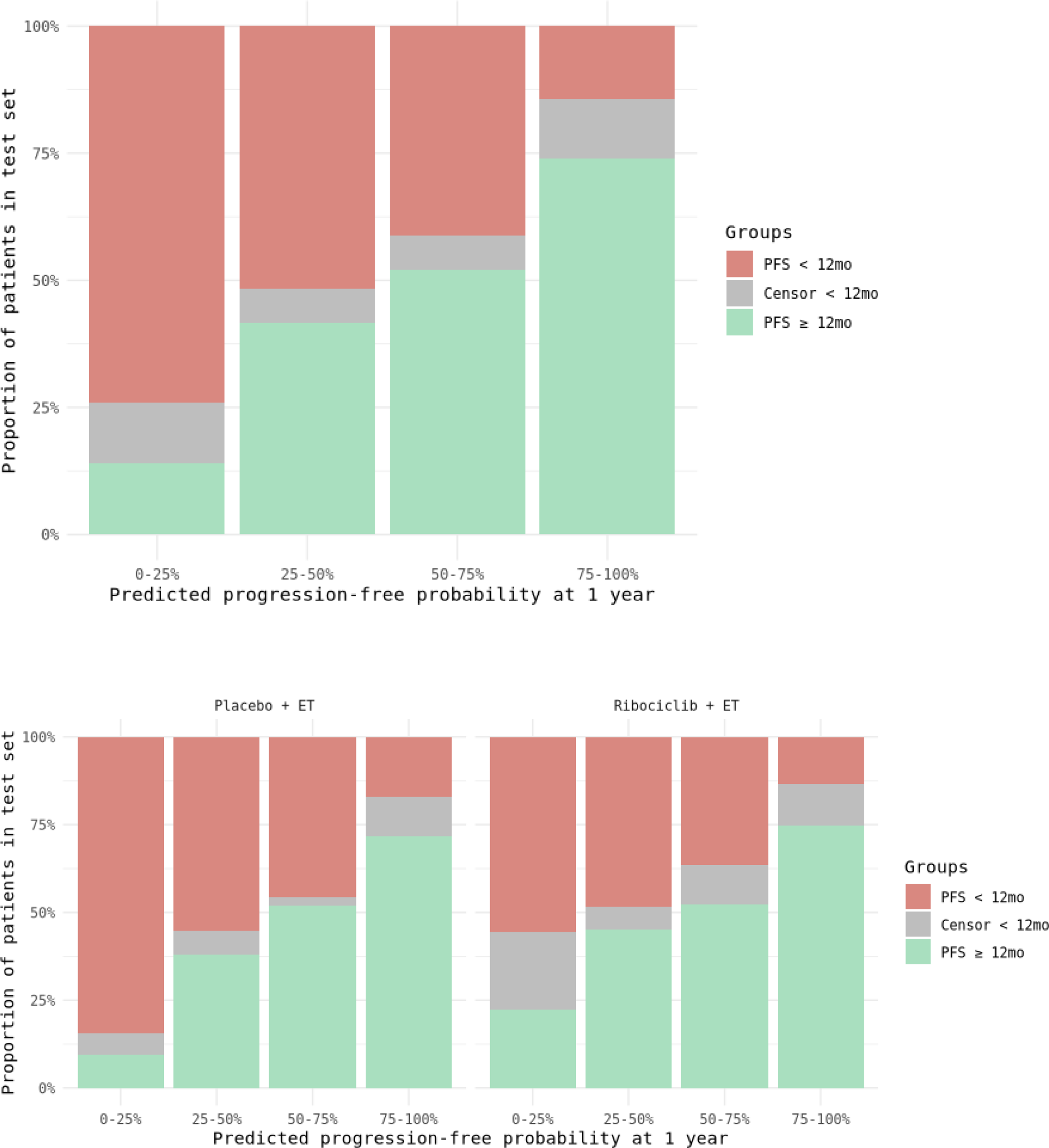
Model-predicted 12-month progression-free survival probability compared with observed outcomes, overall and by treatment arms. ET, endocrine therapy; mo, months; PFS, progression-free survival.

## Discussion

In this investigation, we developed and evaluated a multimodal prognostic modeling approach using pooled phase III MONALEESA trial data, suggesting improved prediction of PFS compared with single-modality approaches. By evaluating multiple analytic strategies across clinical, ctDNA, and gene expression modalities, we found that models integrating biomarker data with clinical information improved prognostic performance compared with single-modality models based on our developmental phase assessments. Both the early fusion deep learning model and the late fusion model achieved superior discrimination, with the late fusion approach demonstrating the more consistent performance compared to the early fusion model in the held-out test set. These findings suggested the value of combining multimodal data to potentially better characterize patient prognosis and provide a foundation for future applications in risk stratification and trial design.

An important methodological consideration in multimodal prognostic modeling is the choice between early and late fusion modeling strategies. Early fusion integrates all available features across modalities into a single model, enabling the learning of complex cross-modal interactions that may reflect underlying biological relationships. However, this approach uses complete multimodal data for all patients and is therefore susceptible to reduced sample size and potential selection bias when data availability differs across modalities. In contrast, late fusion combines predictions from modality-specific models, allowing each data modality to contribute independently while preserving a larger effective sample size. Our analyses using the late fusion approach underscore the importance of integrating complementary data modalities, such as biomarker and clinical data, to better characterize patient prognosis in ABC. While clinical variables may capture well-established prognostic factors such as disease burden and prior therapies, they do not completely reflect underlying tumor biology. In this study, incorporating gene expression and ctDNA data provided deeper biological characterization and improved risk stratification. Although biomarker data generation can yield smaller data sets due to limited tumor material or assay failures, our multimodal modeling demonstrates that biomarker data adds value beyond clinical data, both in terms of prediction and biological insight. While early fusion may be advantageous when complete multimodal data are available, late fusion may be better suited for clinical trial and real-world settings where data completeness varies across modalities with the downside that it may not capture the complex biological interactions between modalities. More complete data sets across modalities may further maximize the benefits of this integrative approach.

In general, covariate adjustment is known to improve the efficiency of clinical trials through more accurate quantification of treatment effects (33, 34). The FDA guidance on covariate adjustment states that “The main focus of the guidance is on the use of prognostic baseline covariates to improve statistical efficiency for estimating and testing treatment effects” (Adjusting for Covariates in Randomized Clinical Trials for Drugs and Biological Products, Section I, 2023) (34). The effective sample size improvement observed in our covariate-adjusted analysis using transformed risk score represents a post-hoc evaluation of the prognostic model and we interpret these scores with caution. The merits of prospective application in clinical trials are a much wider topic beyond the scope of this paper.

Prognostic modeling approaches of this nature have been explored in the context of prognostic enrichment strategies, and the findings from this study may contribute to that body of evidence (34). Focusing on patient cohorts that have higher progression risk allows populations with the greatest unmet need to be targeted. Importantly, our final prognostic model demonstrated strong separation between low- and high-risk patient groups, with consistent performance across clinically relevant subgroups. Estimated 12-month PFS probabilities from the final model provided a clinically relevant metric, and a good correlation between predicted and observed outcomes was observed. If appropriately validated, such a metric may offer a more directly interpretable measure of patient prognosis than relative risk rankings and could serve as one input among others in research contexts aimed at identifying patients at higher risk of early progression. Such a risk model may offer one approach to help characterize patients with less aggressive cancer types who may have higher underlying risk, and to identify patients at higher risk of a progression event within 12 months, a finding that may be explored in future prospective studies.

Clinical and biological factors have traditionally underpinned prognostic assessment in oncology, many of which were also identified in this study (10). For example, liver involvement, histological grade, *de novo* disease, prior ET, time to progression on ET, and Eastern Cooperative Oncology Group performance status were found as well-established risk-factors. Higher levels of ctDNA in the circulation (purity) and higher number of driver mutations have been linked with poorer prognosis in various cancers, including breast cancer (35). Higher *STC2* expression has been associated with improved relapse-free survival in endocrine receptor-positive, particularly luminal A subtype, consistent with its responsiveness to estrogen activation and links to favorable prognosis (36, 37). *RBBP8* (CtIP) is a *BRCA1*-interacting DNA damage repair factor, and reduced expression has been associated with higher grade, hormone receptor negative, basal-like subtype and more aggressive breast cancer, supporting our observation that higher expression is linked with longer PFS (38). *FBXO5* (*EMI1*) regulates the cell cycle and promotes tumor growth and survival, frequently overexpressed in more aggressive types of breast cancer and elevated *FBXO5* expression has been linked to poorer prognosis, consistent with our observation that lower *FBXO5* expression is associated with longer PFS (39, 40). Interestingly, *SLC39A6* (also known as LIV-1), a zinc transporter protein, was identified as a factor. *SLC39A6* is often overexpressed in estrogen receptor-positive (ER+) luminal A subtype, and its overexpression is linked with a better prognosis, which is consistent with our findings that higher *SLC39A6* gene expression is associated with better PFS (41). *SLC39A6* has been a key target for antibody-drug conjugates in ABC treatments (41).

Several newly identified or novel factors were also detected in this study, and validation would be warranted in additional data sets. For example, *DLGAP5* was recently revealed to be highly expressed in more aggressive type of breast cancer and as a potential biomarker which may impact cell proliferation, migration, invasion, cell cycle, and breast cancer development, findings that are supported by our study (42). Although our results are encouraging, with good correlation between model-predicted and observed outcomes, further work and refinement are warranted. Notably, our clinical data set only contains RECIST-based measurements while using higher order radiomics features from whole images may provide additional prognostic information beyond the summary extracted based on RECIST (43). Additionally, machine learning approaches provide the most benefit when there are more samples than features; however, as is common with multiomics data sets, there were fewer samples than features in this study. Finally, it is important to note that while we identified features related to patient prognosis, it remains unknown which features were causal, and which only correlated with outcome. Machine learning methods are increasingly being used to infer causal features from oncology omics data sets and have the potential to better inform treatment decisions and identify potential therapeutic targets.

Our analyses had several limitations. Although the models are intended to be prognostic, the inclusion of randomized treatment arm as a feature introduced potential ambiguity between prognostic and predictive effects in the deep learning models. Consequently, the risk scores and their feature importance are to be interpreted with caution. The models were developed and evaluated using data from randomized phase III clinical trials with defined inclusion and exclusion criteria, which may enroll patients with different characteristics than those treated in routine clinical practice. As a result, progression patterns and prognostic relationships observed in this analysis may not fully reflect those in broader real-world populations. External validation in independent clinical trial data sets and real-world cohorts would be an important step toward establishing the robustness and clinical utility of the proposed prognostic approach.

Looking ahead, the general trend in oncology and biomedical research points to an increasing availability of multimodal data—including imaging, genomics, and digital pathology data. Advances in artificial intelligence and machine learning are rapidly enhancing our ability to process and integrate these complex data sets. As more data modalities become routinely accessible and AI techniques continue to evolve, this multimodal approach may serve as one model for future exploratory research, with the potential to inform risk stratification approaches in clinical trial contexts.

In conclusion, this multimodal prognostic analysis of pooled MONALEESA trial data demonstrates that integrating clinical variables with ctDNA and gene expression data using machine learning approaches may meaningfully improve prediction of PFS in HR+/HER2− ABC. These findings support further exploration of multimodal prognostic models as potential tools for risk stratification and clinical trial design in HR+/HER2− ABC.

## Supporting information

Supplementary Materials - List of IEC/IRBs

## Data Availability

Novartis is committed to sharing with qualified external researchers, access to patient-level data and supporting clinical documents from eligible studies. These requests are reviewed and approved by an independent review panel on the basis of scientific merit. All data provided is anonymized to respect the privacy of patients who have participated in the trial in line with applicable laws and regulations. This trial data availability is according to the criteria and process described on www.clinicalstudydatarequest.com

## REFERENCE

1. Giaquinto AN, Sung H, Newman LA, Freedman RA, Smith RA, Star J, et al. Breast cancer statistics 2024. CA Cancer J Clin 2024;74(6):477–95.

2. Siegel RL, Kratzer TB, Wagle NS, Sung H, Jemal A. Cancer statistics, 2026. CA Cancer J Clin 2026;76:e70043.

3. Huppert LA, Gumusay O, Idossa D, Rugo HS. Systemic therapy for hormone receptor-positive/human epidermal growth factor receptor 2-negative early stage and metastatic breast cancer. CA Cancer J Clin 2023;73(5):480–515.

4. Gradishar WJ, Moran MS, Abraham J, Abramson V, Aft R, Agnese D, et al. NCCN Guidelines® Insights: Breast Cancer, Version 5.2025. J Natl Compr Canc Netw 2025;23(11):426–36.

5. Hortobagyi GN, Stemmer SM, Burris HA, Yap YS, Sonke GS, Hart L, et al. Overall survival with ribociclib plus letrozole in advanced breast cancer. N Engl J Med 2022;386:942–50.

6. Slamon DJ, Neven P, Chia S, Fasching PA, De Laurentiis M, Im SA, et al. Overall survival with ribociclib plus fulvestrant in advanced breast cancer. N Engl J Med 2020;382:514–24.

7. Im SA, Lu YS, Bardia A, Harbeck N, Colleoni M, Franke F, et al. Overall survival with ribociclib plus endocrine therapy in breast cancer. N Engl J Med 2019;381:307–16.

8. Novartis Pharmaceuticals Corporation. KISQALI® (ribociclib) tablets, for oral use: Prescribing information. U.S. Food and Drug Administration; 2024. Revised September 2024. Available from: https://www.accessdata.fda.gov/drugsatfda_docs/label/2024/209092s018lbl.pdf.

9. Shah A, Bloomquist E, Tang S, Fu W, Bi Y, Liu Q, et al. FDA approval: ribociclib for the treatment of postmenopausal women with hormone receptor-positive, HER2-negative advanced or metastatic breast cancer. Clin Cancer Res 2018;24(13):2999–3004.

10. Cuyún Carter G, Mohanty M, Stenger K, Morato Guimaraes C, Singuru S, Basa P, et al. Prognostic factors in hormone receptor-positive/human epidermal growth factor receptor 2-negative (HR+/HER2-) advanced breast cancer: a systematic literature review. Cancer Manag Res 2021;13:6537–66.

11. Kernan WN, Viscoli CM, Makuch RW, Brass LM, Horwitz RI. Stratified randomization for clinical trials. J Clin Epidemiol 1999;52:19–26.

12. Fathi Kazerooni A, Saxena S, Toorens E, Tu D, Bashyam V, Akbari H, et al. Clinical measures, radiomics, and genomics offer synergistic value in AI-based prediction of overall survival in patients with glioblastoma. Sci Rep 2022;12:8784.

13. Nassar AH, Mouw KW, Jegede O, Shinagare AB, Kim J, Liu CJ, et al. A model combining clinical and genomic factors to predict response to PD-1/PD-L1 blockade in advanced urothelial carcinoma. Br J Cancer 2020;122:555–63.

14. Tong D, Tian Y, Zhou T, Ye Q, Li J, Ding K, et al. Improving prediction performance of colon cancer prognosis based on the integration of clinical and multi-omics data. BMC Med Inform Decis Mak 2020;20:22.

15. Deng Y, Song S, Zhao H, Yang Y, Lu S, Li X. Artificial intelligence-based evaluation of prognostic benefits from immunotherapy plus targeted therapy with or without radiotherapy or TACE in advanced hepatocellular carcinoma. Front Oncol 2025;15:1694565.

16. Esteva A, Feng J, van der Wal D, Huang SC, Simko JP, DeVries S, et al. Prostate cancer therapy personalization via multi-modal deep learning on randomized phase III clinical trials. NPJ Digit Med 2022;5:71.

17. Jiang N, Xiong X, Chen X, Feng M, Guo Y, Hu C. Machine learning and deep learning to improve overall survival prediction in cervical cancer patients. Transl Cancer Res 2025;14:3057–68.

18. Yalçıner M, Erdat EC, Kavak EE, Utkan G. Development and validation of an AI-augmented deep learning model for survival prediction in de novo metastatic colorectal cancer. Discov Oncol 2025;16:2126.

19. Coroller T, Sahiner B, Amatya A, Gossmann A, Karagiannis K, Moloney C, et al. Methodology for good machine learning with multi-omics data. Clin Pharmacol Ther 2024;115(4):745–57.

20. Yardley DA. MONALEESA clinical program: a review of ribociclib use in different clinical settings. Future Oncol 2019;15:2673–86.

21. Hortobagyi GN, Stemmer SM, Burris HA, Yap YS, Sonke GS, Paluch-Shimon S, et al. Ribociclib as first-line therapy for HR-positive, advanced breast cancer. N Engl J Med 2016;375:1738–48.

22. Slamon DJ, Neven P, Chia S, Fasching PA, De Laurentiis M, Im SA, et al. Phase III randomized study of ribociclib and fulvestrant in hormone receptor-positive, human epidermal growth factor receptor 2-negative advanced breast cancer: MONALEESA-3. J Clin Oncol 2018;36:2465–72.

23. Tripathy D, Im SA, Colleoni M, Franke F, Bardia A, Harbeck N, et al. Ribociclib plus endocrine therapy for premenopausal women with hormone-receptor-positive, advanced breast cancer (MONALEESA-7): a randomised phase 3 trial. Lancet Oncol 2018;19:904–15.

24. Coroller T, Sahiner B, Amatya A, Gossmann A, Karagiannis K, Samala RK, et al. Multi-omics investigation on the prognostic and predictive factors in metastatic breast cancer using data from phase III ribociclib clinical trials: a statistical and machine learning analysis plan. medRxiv 2023. Available from: 10.1101/2023.08.30.23294367.

25. Hsu JC, Wu M, Kim C, Vora B, Lien YTK, Jindal A, et al. Applications of advanced natural language processing for clinical pharmacology. Clin Pharmacol Ther 2024;115:786–94.

26. Meinshausen N, Bühlmann P. Stability Selection. J R Stat Soc Ser B Stat Methodol 2010;72:417–73.

27. Monod M, Krusche P, Cao Q, Sahiner B, Petrick N, Ohlssen D, et al. TorchSurv: a lightweight package for deep survival analysis. The Journal of Open Source Software 2024;9:7341.

28. US Food and Drug Administration. TorchSurv: deep learning tools for survival analysis (RST24AI17.01). [cited 2026 Jan 5]. Available from: https://cdrh-rst.fda.gov/torchsurv-deep-learning-tools-survival-analysis.

29. Gardner MW, Dorling SR. Artificial neural networks (the multilayer perceptron)—a review of applications in the atmospheric sciences. Atmos Environ 1998;32:2627–36.

30. Rahman MS, Ambler G, Choodari-Oskooei B, Omar RZ. Review and evaluation of performance measures for survival prediction models in external validation settings. BMC Med Res Methodol 2017;17:60.

31. Ye T, Shao J, Yi Y. Covariate-adjusted log-rank test: guaranteed efficiency gain and universal applicability. Biometrika 2024;111:691–705.

32. Lundberg SM, Lee S. A Unified Approach to Interpreting Model Predictions. Neural Information Processing Systems 2017.

33. Benkeser D, Díaz I, Luedtke A, Segal J, Scharfstein D, Rosenblum M. Improving precision and power in randomized trials for COVID-19 treatments using covariate adjustment, for binary, ordinal, and time-to-event outcomes. Biometrics 2021;77:1467–81.

34. U.S. Food and Drug Administration. Guidance document: adjusting for covariates in randomized clinical trials for drugs and biological products: guidance document: for industry. Final guidance. May 2023. Published 2023. Accessed January 5, 2025. https://www.fda.gov/regulatory-information/search-fda-guidance-documents/.

35. Mamann A, Pradat Y, Bidard FC, Delaloge S, Cabel L, Faull I, et al. Prognostic significance of early on-treatment evolution of circulating tumor DNA in advanced ER-positive/HER2-negative breast cancer. Ann Oncol 2025;36:1342–55.

36. Qie S, Sang N. Stanniocalcin 2 (STC2): a universal tumour biomarker and a potential therapeutical target. J Exp Clin Cancer Res 2022;41:161.

37. Muñoz JP, Lampe-Huenul N. Predictive value of STC2 gene expression in chemotherapy response in breast cancer. Pharmaceuticals (Basel) 2025;18:235.

38. Soria-Bretones I, Sáez C, Ruíz-Borrego M, Japon MA, Huertas P. Prognostic value of CtIP/RBBP8 expression in breast cancer. Cancer Med 2013;2:774–83.

39. Vaidyanathan S, Cato K, Tang L, Pavey S, Haass NK, Gabrielli BG, et al. In vivo overexpression of Emi1 promotes chromosome instability and tumorigenesis. Oncogene 2016;35:5446–55.

40. Liu P, Wang X, Pan L, Han B, He Z. Prognostic significance and immunological role of FBXO5 in human cancers: a systematic pan-cancer analysis. Front Immunol 2022;13:901784.

41. Althobiti M, El-Sharawy KA, Joseph C, Aleskandarany M, Toss MS, Green AR, et al. Oestrogen-regulated protein SLC39A6: a biomarker of good prognosis in luminal breast cancer. Breast Cancer Res Treat 2021;189:621–30.

42. Li Y, Wei J, Sun Y, Zhou W, Ma X, Guo J, et al. DLGAP5 regulates the proliferation, migration, invasion, and cell cycle of breast cancer cells via the JAK2/STAT3 signaling axis. Int J Mol Sci 2023;24:15819.

43. Khorrami M, Viswanathan VS, Reddy P, Braman N, Kunte S, Gupta A, et al. Radiomic predicts early response to CDK4/6 inhibitors in hormone receptor positive metastatic breast cancer. NPJ Breast Cancer 2023;9:67.

