## Supplementary Materials for "Multimodal Machine Learning–Based Clinical and Genomic Prognostic Factors for Progression-Free Survival in Metastatic Breast Cancer Using Phase III MONALEESA Trials"

Additional details on data collection and analytical methods

#### Data collection - circulating tumor DNA

Cell-free DNA was extracted from plasma samples collected prior to treatment and sequenced using an Illumina HiSeq instrument targeting the exonic regions of 558 genes implicated in cancer. Single-nucleotide variants and indels were identified using MuTect (Broad Institute, Cambridge, Massachusetts) and Pindel (1, 2), respectively, and copy number alterations were assessed using PureCN (3). Detailed methods regarding removal of germline variants and artefacts have been described previously (4). Somatic mutations due to clonal hematopoiesis of indeterminate potential were identified and removed (4, 5). Synonymous mutations and synonymous indels were also removed. Tumor mutational burden was estimated as the number of somatic mutations per megabase. The machine learning modeling excluded sparse circulating tumor DNA (ctDNA) features with a frequency <3% which would generally lead to less than 10 mutated patients in one or both arms.

#### Data collection - gene expression

Formalin-fixed paraffin-embedded tumor samples (tumor block or slides) were collected in each of the trials. Gene expression data in tumor samples with >10% tumor content was generated using a custom CodeSet gene panel (781 genes) and the nCounter platform from Nanostring Technologies. Housekeeping gene normalization was performed on the NanoString raw counts.

#### Gene expression data - investigation of intervening line of therapy

Archival tumor biopsies were collected prior to randomization, with the time frame for these biopsies ranging from a few months to several years. Considering that tumors can undergo significant evolution over time, particularly in response to treatments, gene expression data were modelled across different patient subsets; these included all patients, patients with no intervening line of therapy between the time of tumor biopsy and randomization, and patients who had received an intervening line of therapy between their tumor biopsy and randomization. The concordance index (C-index) for the model that included patients who had an intervening line of therapy was lower compared to the model excluding such patients. However, the model that encompassed all patients demonstrated a C-index comparable to the model excluding patients with an intervening line of therapy and exhibited lesser variance in the C-index distribution (data not shown). Based on these observations and recognizing the advantage of utilizing a larger sample size, we included all patients with available gene expression data regardless of intervening line of therapy in subsequent modeling. To account for the potential biological impact of an intervening line of therapy, we included an additional feature representing this factor in the gene expression model.

#### Feature engineering - imputation and standardization

The median-based imputation approach was utilized for numerical variables and mode-based imputation for categorical variables. To prevent data leakage, the imputation of missing variables for any patient records was carried out using only the training data set. Specifically, when imputing a single patient record from the training super or testing set, the median/mode-based imputation was applied utilizing the entire training set along with the individual patient record from the training super or testing set.

All categorical features were transformed into dummy variables using one-hot encoding for the deep learning model. All features were then normalized to lie within the range [0, 1] (scaling was based on the training set for consistency across all data sets) to avoid any poorly adjusted gradients during the modelling phase. Importantly, when using the scale of the training set for normalization, the normalization of features in the validation and test sets may not fall perfectly within the [0, 1] range. This may occur because the feature distributions in the validation and test sets can differ slightly from those in the training set.

Since treatment arm was not a patient characteristic but instead assigned during trial randomization, it was considered as a feature for each clinical, ctDNA, and gene expression individual modality models.

#### Analytical methods - rational for feature pre-selection

Due to the limited data availability of each modality compared to the number of candidate features, further splitting the data set directly into training and validation for feature selection may have caused unreliable performance assessments. Furthermore, the features selected using such a method may not be representative of variables for the entire data set due to small sample sizes. As a result, we considered a prefiltering step using univariate Cox regression on all available training sets (6).

#### Analytical methods - hyperparameter tuning

To optimize machine learning model performance, a Bayesian-inspired hyperparameter tuning using the Optuna framework was employed (7). Optuna is an efficient, automatic hyperparameter optimization library that leverages sequential model-based optimization strategies combined with pruning techniques to reduce unnecessary evaluations.

We focused on optimizing the CoxnetSurvivalAnalysis and Random Survival Forest (RSF) model. For CoxnetSurvivalAnalysis, two key hyperparameters were tuned:

- α_min_ratio: controlling the minimum value of the regularization strength (searched between 0.01 and 0.2), and
- l1_ratio: balancing the l1 and l2 penalties (searched between 0.1 and 1.0).

Three hyper-parameters for RSF were assessed:

- n_estimators: number of trees in the forest (searched between 50 and 400),
- max_depth: maximum depth of each tree (searched between 5 and 15), and
- min_samples_leaf: minimum number of samples per terminal node (searched between 5 and 15).

For deep learning, three sets of tuning parameters were considered for the grid search implementation, with learning rate set at 0.005 and the batch side for the stochastic gradient descent is set at 96:

- Number of hidden layers (1, 2, 3),
- Number of nodes in each layer (16, 32, 48), and
- Percentage of dropout in each layer (0%, 20%, 40%).

Each optimization run was conducted for 250 trials. Optuna’s Median Pruner was used to terminate underperforming trials early. To account for variability due to random train-validation splits, the optimization procedure was repeated 50 times with different random seeds. For each repetition, a new study was created, and the best trial parameters and validation performance were recorded. This approach provided a distribution of optimal hyperparameters and allowed assessment of the stability of solutions across random splits.

For the deep learning models, the hyperparameter of interest was the number of hidden layers, nodes per hidden layer, and learning and dropout rate.

### Supplementary Tables and Figures

**Table S1.** Sample size available and features for each data modality.

| **Data availability** | **Sample size** | **Number of candidate features** | **Number of features after stable feature selection** |
| --- | --- | --- | --- |
| **Training sets** |  |  |  |
| Clinical | 1,113 | 72 | 25 |
| ctDNA | 833 | 32 | 27 |
| Gene expression | 470 | 782 | 80 |
| Early fusion set^a^ | 336 | 884 | 135 |
| **Evaluation set** | 242 | 884 | 135 |
| **Testing set** | 382 | 884 | 135 |

^a^Early fusion future set consisted of patients with non-missing data across clinical, ctDNA, and gene expression modalities. CtDNA, circulating tumor DNA.

**Figure S1.** Prognostic performance of the multimodal risk score across clinically relevant subgroups.

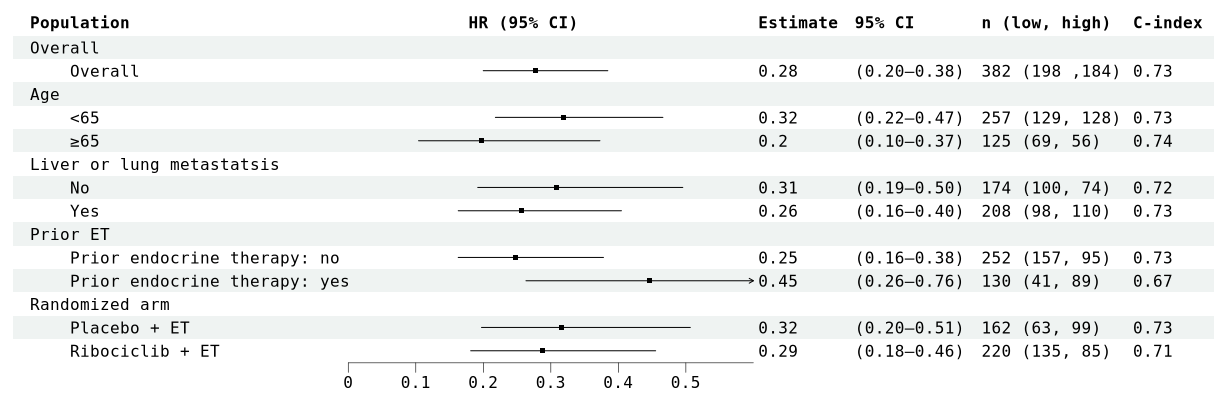

n (low, high) denotes sample size and number of low-risk and high-risk patients within each subgroup. C-index was based on the evaluation on test set. C-index, concordance index; CI, confidence interval; ET, endocrine therapy; HR, hazard ratio; PBO, placebo; TRT, treatment.

**Figure S2. SHAP plots of the individual models used in late fusion, evaluated on test set.**

A. Clinical model using deep learning (top 50 clinical features are shown).

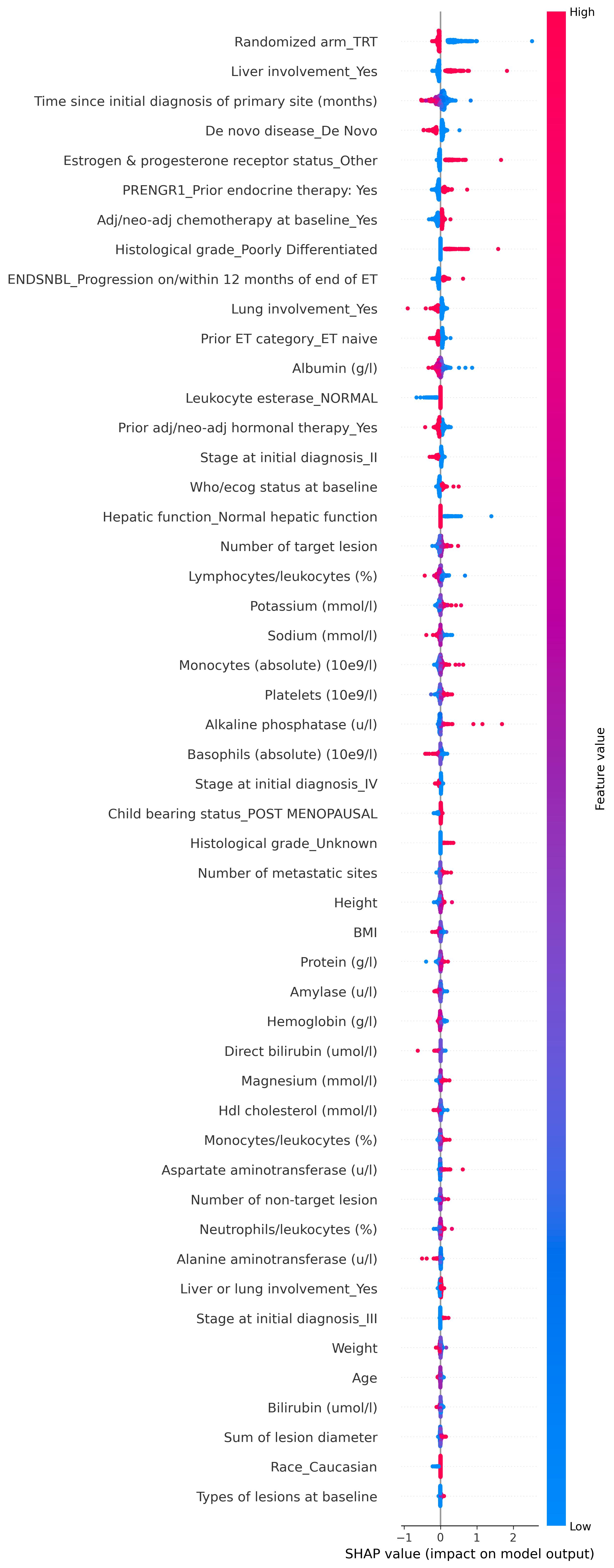

B. ctDNA model using elastic net.

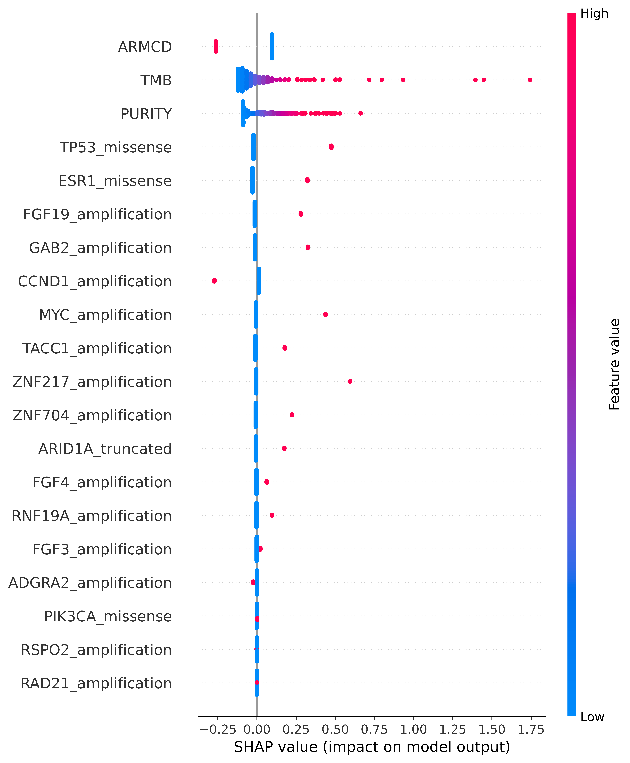

C. Gene expression model using elastic net
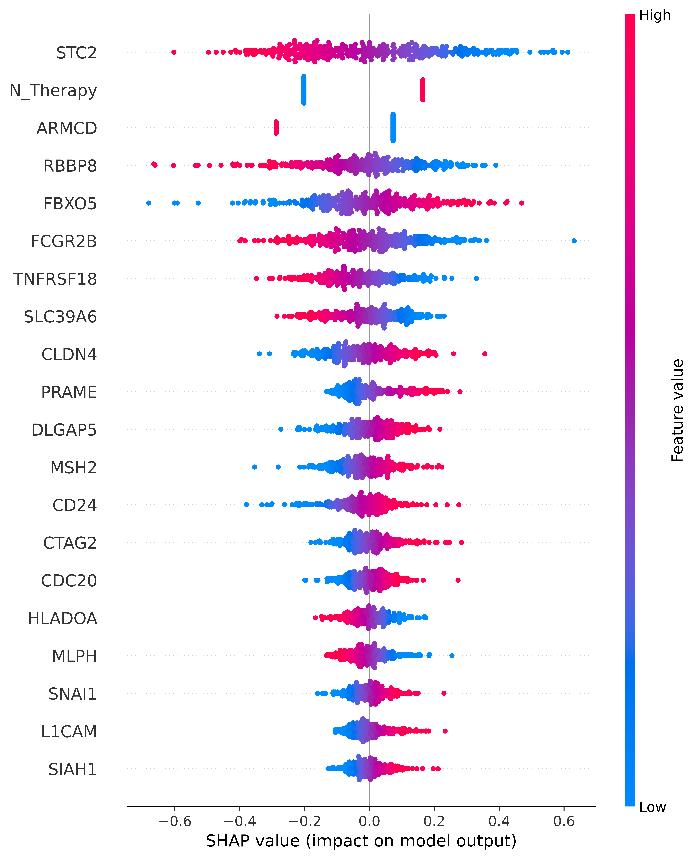
.

ADGRA2, adhesion G protein‑coupled receptor A2; adj, adjuvant; ARID1A, at‑rich interaction domain 1a; ARMCD, armadillo repeat containing domain; BMI, body mass index; CCND1, cyclin D1; CD24, cluster of differentiation 24; CDC20, cell division cycle 20; CLDN4, claudin 4; CTAG2, cancer/testis antigen 2; ctDNA, circulating tumor DNA; DLGAP5, discs large associated protein 5; ESR1, estrogen receptor 1; ET, endocrine therapy; FBXO5, F box protein 5; FCGR2B, Fc gamma receptor IIB; FGF19, fibroblast growth factor 19; FGF3, fibroblast growth factor 3; FGF4, fibroblast growth factor 4; GAB2, GRB2‑associated binding protein 2; HDL, high-density lipoprotein; HLADOA, HLA class II histocompatibility antigen DO alpha chain; L1CAM, L1 cell adhesion molecule; MLPH, melanophilin; MSH2, MutS homolog 2; MYC, Myc proto‑oncogene; N_therapy, number of therapies; PIK3CA, phosphatidylinositol‑4,5‑bisphosphate 3‑kinase catalytic subunit alpha; PRAME, preferentially expressed antigen in melanoma; PURITY, ctDNA purity; RAD21, RAD21 cohesin complex component; RBBP8, retinoblastoma binding protein 8; RNF19A, ring finger protein 19a; RSPO2, R‑spondin 2; SHAP, Shapley additive explanation; SIAH1, SIAH e3 ubiquitin protein ligase 1; SLC39A6, solute carrier family 39 member 6; SNAI1, snail family transcriptional repressor 1; STC2, stanniocalcin 2; TACC1, transforming acidic coiled‑coil containing protein 1; TMB, tumor mutational burden; TNFRSF18, tumor necrosis factor receptor superfamily member 18; TP53, tumor protein p53; WHO/ECOG, World Health Organization/Eastern Cooperative Oncology Group; ZNF217, zinc finger protein 217; ZNF704, zinc finger protein 704.

**REFERENCES**

1.	Ye K, Schulz MH, Long Q, Apweiler R, Ning Z. Pindel: a pattern growth approach to detect break points of large deletions and medium sized insertions from paired-end short reads. *Bioinformatics* **2009**;25:2865–71.

2.	Cibulskis K, Lawrence MS, Carter SL, Sivachenko A, Jaffe D, Sougnez C*, et al.* Sensitive detection of somatic point mutations in impure and heterogeneous cancer samples. *Nat Biotechnol* **2013**;31:213–9.

3.	Riester M, Singh AP, Brannon AR, Yu K, Campbell CD, Chiang DY*, et al.* PureCN: copy number calling and SNV classification using targeted short read sequencing. *Source Code Biol Med* **2016**;11:13.

4.	André F, Su F, Solovieff N, Hortobagyi G, Chia S, Neven P*, et al.* Pooled ctDNA analysis of MONALEESA phase III advanced breast cancer trials. *Ann Oncol* **2023**;34:1003–14.

5.	Fairchild L, Whalen J, D'Aco K, Wu J, Gustafson CB, Solovieff N*, et al.* Clonal hematopoiesis detection in patients with cancer using cell-free DNA sequencing. *Sci Transl Med* **2023**;15:eabm8729.

6.	Meinshausen N, Bühlmann P. Stability selection. *J R Stat Soc Ser B Stat Methodol* **2010**;72:417–73.

7.	Akiba T, Sano S, Yanase T, Ohta T, Koyama M. Optuna: a next-generation hyperparameter optimization framework. In: KDD '19: Proceedings of the 25th ACM SIGKDD International Conference on Knowledge Discovery & Data Mining. New York: Association for Computing Machinery; 2019. p. 2623–31.
