## Supplementary Materials - List of IEC/IRBs for "Multimodal Machine Learning–Based Clinical and Genomic Prognostic Factors for Progression-Free Survival in Metastatic Breast Cancer Using Phase III MONALEESA Trials"

### **List of Independent Ethics Committees (IECs) or Institutional Review Boards (IRBs)**

Table 1. List of Independent Ethics Committees (IEC) or Institutional Review Boards (IRB) by study center with ethical approvals for MONALEESA 2

| **Center No.** | **Ethics Committee or Institutional Review Board** | **Department / Organization** | **Address** |
| --- | --- | --- | --- |
| 4100 | Comite de Etica en Investigación Clinica. (CEIC) | CABA | CABA, Buenos Aires, C1117ABK, Argentina |
| 4101 | Comité Institucional de Ética de la Investigación en salud Clínica Colombo | Cordoba | Cordoba, Cordoba, 5000, Argentina |
| 4102 | Comite de Etica en Investigación Clinica. (CEIC) | CABA | CABA, Buenos Aires, C1117ABK, Argentina |
| 1001 | Bellberry Limited |  | South Adelaide, SA, 5063, Australia |
| 1002 | Bellberry Limited |  | South Adelaide, SA, 5063, Australia |
| 1003 | Bellberry Limited |  | South Adelaide, SA, 5063, Australia |
| 2500 | Ethikkommission der Stadt Wien |  | Wien,1030, Austria |
| 2502 | Ethikkommission für das Bundesland Salzburg |  | Salzburg, 25010, Austria |
| 2503 | Ethikkommission der Medizinischen Universität Wien |  | Borschkegasse 8b/E06, Vienna, A-1090, Austria |
| 2600 | AZ Nikolaas | EC | Sint-Niklaas, 9100, Belgium |
| 2601 | Clinique et maternité Sainte-Elisabethe | Ethics committee | Namur, 5000, Belgium |
| 2602 | Jessa | EC | Hasselt, 3500, Belgium |
| 2603 | Sint Augustinus | EC | Wilrijk, 2610, Belgium |
| 2604 | Ethische commissie onderzoek UZ / KU Leuven |  | Leuven, 3000, Belgium |
| 4200 | Comitê de Ética em Pesquisa do Hospital Pérola Byington |  | São Paulo, São Paulo, 01317-000, Brazil |
| 4202 | Comite de Etica em Pesquisa do Hospital das Clinicas da Faculdade de Medicina de Ribeirao Preto FMRP-USP |  | Ribeirao Preto, SP,14048-900, Brazil |
| 4203 | Comitê de Ética em Pesquisa do Hospital das Clinicas da Faculdade de Medicina da USP HCFMUSP |  | Sao Paulo, SP, 05403010, Brazil |
| 6001 | OCREB | MaRs Centre South Tower, Suite 800 ,101 College Street | Toronto, Ontario, M5G 0A3, Canada |
| 6002 | OCREB | MaRs Centre, South Tower, Suite 800 101 College Street | Toronto, Ontario, M5G 0A3, Canada |
| 6003 | OCREB | MARS Center suite 510, 661 University Avenue | Toronto, ON, M5G 0A3, Canada |
| 6004 | Fraser Health Research Ethics board | FHA evaluation and research services | Surrey, BC, V3T 0H1, Canada |
| 6005 | Comité d'éthique de la recherche CHU de Québec - Université Laval | 10, rue de l'Espinay Édifice D, 7 étage | Quebec, Quebec, G1L 3L5, Canada |
| 6006 | CÉR du CHUM | Tour Vige, 900 rue St-Denis, 3e étage | Montreal, Qc, H2X 0A9, Canada |
| 6007 | Ontario Cancer Research Ethics Board | MARS Centre, South Tower, suite 800, 101 College Street | Toronto, ON, M5G 0A3, Canada |
| 6008 | Nova Scotia Health Authority Research Ethics Board | Centre for Clinical Research, Room 118, 5790 University Avenue | Halifax, NS, B3H 1V7, Canada |
| 2700 | Etická komise FN s LF UP Olomouc |  | Olomouc, 77520, Czech Republic |
| 2701 | Etická komise MOU |  | Brno, 656 53, Czech Republic |
| 2703 | Eticka komise nemocnice Liberec |  | Liberec, 46063,Czech Republic |
| 2706 | Multicentricka eticka komise Fakultni nemocnice Brno |  | Brno, 625 00, Czech Republic |
| 2800 | De videnskabsetiske Komitéer for Region Syddanmark | Regionshuset, Damhaven 12 | Vejle, 7100, Denmark |
| 2801 | De videnskabsetiske Komitéer for Region Syddanmark | Regionshuset, Damhaven 12 | Vejle, 7100, Denmark |
| 2802 | De videnskabsetiske Komitéer for Region Syddanmark | Regionshuset, Damhaven 12 | Vejle, 7100, Denmark |
| 2805 | De videnskabsetiske Komitéer for Region Syddanmark | Regionshuset, Damhaven 12 | Vejle, 7100, Denmark |
| 2900 | VSSHP | Ethics committee | Turku, 20520, Finland |
| 2903 | VSSHP | Ethics committee | Turku, 20520, Finland |
| 3000 | CPP OUEST IV | CHU de Nantes - 5 allée de l'ile gloriette | Nantes Cedex 1, 44093, France |
| 3001 | CPP OUEST IV | CHU de Nantes - 5 allée de l'ile gloriette | Nantes Cedex 1, 44093, France |
| 3002 | CPP OUEST IV | CHU de Nantes - 5 allée de l'ile gloriette | Nantes Cedex 1, 44093, France |
| 3003 | CPP OUEST IV | CHU de Nantes - 5 allée de l'ile gloriette | Nantes Cedex 1, 44093, France |
| 3004 | CPP OUEST IV | CHU de Nantes - 5 allée de l'ile gloriette | Nantes Cedex 1,44093, France |
| 3006 | CPP OUEST IV | CHU de Nantes - 5 allée de l'ile gloriette | Nantes Cedex 1,44093, France |
| 3007 | CPP OUEST IV | CHU de Nantes - 5 allée de l'ile gloriette | Nantes Cedex 1, 44093, France |
| 3008 | CPP OUEST IV | CHU de Nantes - 5 allée de l'ile gloriette | Nantes Cedex 1, 44093, France |
| 3009 | CPP OUEST IV | CHU de Nantes - 5 allée de l'ile gloriette | Nantes Cedex 1,44093, France |
| 3011 | CPP OUEST IV | CHU de Nantes - 5 allée de l'ile gloriette | Nantes Cedex 1, 44093, France |
| 3012 | CPP OUEST IV | CHU de Nantes - 5 allée de l'ile gloriette | Nantes Cedex 1, 44093, France |
| 3014 | CPP OUEST IV | CHU de Nantes - 5 allée de l'ile gloriette | Nantes Cedex 1, 44093, France |
| 3100 | Universtität Ulm | Ethikkommission | Ulm, 89081, Germany |
| 3101 | Universtität Ulm | Ethikkommission | Ulm, 89081, Germany |
| 3103 | Universtität Ulm | Ethikkommission | Ulm, 89081, Germany |
| 3105 | Universtität Ulm | Ethikkommission | Ulm, 89081, Germany |
| 3106 | Universtität Ulm | Ethikkommission | Ulm, 89081, Germany |
| 3107 | Universtität Ulm | Ethikkommission | Ulm, 89081, Germany |
| 3109 | Universtität Ulm | Ethikkommission | Ulm, 89081, Germany |
| 3110 | Universtität Ulm | Ethikkommission | Ulm, 89081, Germany |
| 3112 | Universtität Ulm | Ethikkommission | Ulm, 89081, Germany |
| 3113 | Universtität Ulm | Ethikkommission | Ulm, 89081, Germany |
| 3114 | Universtität Ulm | Ethikkommission | Ulm, 89081, Germany |
| 3115 | Universtität Ulm | Ethikkommission | Ulm, 89081, Germany |
| 3116 | Universtität Ulm | Ethikkommission | Ulm, 89081, Germany |
| 3117 | Universtität Ulm | Ethikkommission | Ulm, 89081, Germany |
| 3118 | Universtität Ulm | Ethikkommission | Ulm, 89081, Germany |
| 3119 | Universtität Ulm | Ethikkommission | Ulm, 89081, Germany |
| 3120 | Universtität Ulm | Ethikkommission | Ulm, 89081, Germany |
| 3121 | Universtität Ulm | Ethikkommission | Ulm, 89081, Germany |
| 3123 | Universtität Ulm | Ethikkommission | Ulm, 89081, Germany |
| 3126 | Universtität Ulm | Ethikkommission | Ulm, 89081, Germany |
| 3201 | Medical Research Council, Ethics Committee for Clinical Pharmacology |  | Budapest, 1051, Hungary |
| 3202 | Medical Research Council, Ethics Committee for Clinical Pharmacology |  | Budapest, 1051, Hungary |
| 3205 | Medical Research Council, Ethics Committee for Clinical Pharmacology |  | Budapest, 1051, Hungary |
| 3207 | Medical Research Council, Ethics Committee for Clinical Pharmacology |  | Budapest, 1051, Hungary |
| 3960 | Clinical Research Ethics Committee of the Cork Teaching Hospital |  | Lancaster Hall,6 Little Hanover, Cork, Ireland |
| 3961 | St Vincent's Healthcare | Ethics and Medical Research Committee | Elm Park,Dublin 4,  Ireland |
| 1300 | The Chaim Sheba Helsinki committee |  | Ramat Gan, 5266202, Israel |
| 1301 | The Tel Aviv Sourasky Helsinki committee |  | 6 Weizman street, 6423906, Israel |
| 1302 | Rabin MC Helsinki | Helsinki | Petah Tikva, 4941492, Israel |
| 3300 | Comitato Etico Regionale Toscana-Area Vasta Nord Ovest - Azienda Ospedaliero Università Pisana di Pisa | Azienda Ospedaliero Unoversitaria Pisana di Pisa | Pisa, 56126, Italy |
| 3301 | IRCCS di Candiolo | Comitato Etico | Candiolo (TO), 10060, Italy |
| 3302 | Comitato Etico Regione Calabria Sezione Area Sud | Azienda Ospedaliera Bianchi-Melacrino-Morelli Palazzo Gangeri | Reggio Calabria, 89128, Italy |
| 3303 | Comitato Etico Delle Aziende Sanitarie Dell'umbria - Segreteria Scientifico Amministrativa Del Ceas Umbria |  | Perugia, 06124, Italy |
| 3304 | CE per la Sperimentazione Clinica (CESC) | Istituto Oncologico Veneto (IOV) | Padova (PD), 35128, Italy |
| 3308 | Comitato Etico Della Provincia Di Brescia C/O Ao Spedali Civili | Asst Degli Spedali Civili Di Brescia | Brescia, 25123, Italy |
| 3314 | Comitato Etico Regionale Della Liguria C/O Irccs Aou San Martino |  | Genova, 16132, Italy |
| 3315 | Comitato Etico Fondazione IRCCS Istituto Nazionale dei Tumori | via Venezian 1 | Milano, 20133, Italy |
| 3316 | CE Indipendente del Centro Riferimento Oncologico | CRO Aviano | Aviano ,33081,Italy |
| 3317 | CE Lazio 1 | CE | Roma, 00152, Italy |
| 3319 | Comitato Etico Interaziendale Della Provincia Di Messina | C/O Aou Policlinico G. Martino | Messina, 98124, Italy |
| 3320 | Comitato Etico Delle Aziende Sanitarie Dell'umbria - Segreteria Scientifico Amministrativa Del Ceas Umbria |  | Perugia, 06124, Italy |
| 3322 | CE IRCCS dell'Istituto Nazionale per lo studio e la cura dei tumori | Fondazione Pascale | Napoli, 80131, Italy |
| 3323 | Comitato Etico Regionale (Cer) Delle Marche C/O Azienda Ospedaliera | Azienda Ospedaliero Universitaria "Ospedali Riuniti Umberto I, G.M. Lancisi, G. Salesi" di Ancona | Ancona, 60126, Italy |
| 3324 | Comitato Etico dell'Università Cattolica del Sacro Cuore | Fondazione Policlinico Gemelli, Largo Gemelli, 8 | Roma, 00168, Italy |
| 3325 | Comitato Etico della Brianza | Ospedale San Gerardo di Monza | Monza, 20900, Italy |
| 1404 | Asan Medical Center | Institutional Review Board | Songpa-gu, Seoul, 05505, Korea, Republic of |
| 1405 | Yonsei University Severance Hospital Institutional Review Board | Human Research Protection Center | Seoul, Seodaemun-gu, 03722, Korea, Republic of |
| 1406 | National Cancer Center | Institutional Review Board | Goyang-si, Gyeonggi-do, 10408, Korea, Republic of |
| 1407 | Seoul National university bundang hospital IRB | IRB | Bundang-gu, Seongnam-si, Gyeonggi-do, 13620, Korea, Republic of |
| 1408 | Seoul National University Hospital IRB | IRB | Seoul, 101, Daehak-ro, Jongno-gu, 03080, Korea, Republic of |
| 1500 | The Ethics Committee of the Hôtel-Dieu de France | Medical science campus of USJ | Blockt E, 166830, Lebanon |
| 1501 | IRB of Hammoud Hospital University Medical center |  | Beirut, 652, Lebanon |
| 1502 | Institutional Review Board American University of Beirut | Biomedical | Beirut ,1107 2020, Lebanon |
| 1503 | IRB of Bellevue Medical Center |  | Beirut, 295, Lebanon |
| 3400 | Antoni van Leeuwenhoek | Raad van Bestuur | Amsterdam, Noord-Holland, 1066 CX, Netherlands |
| 3401 | Maastricht UMC+ | Raad van Bestuur | Maastricht, 6229 EV, Netherlands |
| 3403 | LUMC | Raad van Bestuur | Leiden, 2333 ZA, Netherlands |
| 3406 | Orbis Medisch Centrum | Directie | Sittard-Geleen, 6162 BG, Netherlands |
| 3409 | Deventer Ziekenhuis | Raad van Bestuur | Deventer, 7416 SE, Netherlands |
| 3411 | Martini Ziekenhuis | Raad van bestuur | Groningen, 9700RM, Netherlands |
| 3414 | Isala Klinieken | Raad van Bestuur | Zwolle, 8025 AB, Netherlands |
| 3416 | Elisabeth-TweeSteden Ziekenhuis | Raad van Bestuur | Tilburg, 5022 GC, Netherlands |
| 3417 | UMCG | Raad van Bestuur | Groningen, 9700 RB, Netherlands |
| 3423 | Noordwest Ziekenhuisgroep | Raad van Bestuur | Alkmaar, 1815 GM, Netherlands |
| 3500 | REK | Sør-Øst | Oslo,0484,Norway |
| 3501 | REK | Sør-Øst | Oslo,0484,Norway |
| 1600 | Arkhangelsk Regional Oncology Center | LEC | bld. 145/1, Obvodniy kanal av., Arkhangelsk, 163045, Russia |
| 1601 | Privolzhskyi Federal Medical Research Center | LEC | bld. 18/1, Verhnevolzhskaya qy., Nizhny Novgorod, 603155, Russia |
| 1602 | Regional Clinical Oncological Dispensary | LEC | bld. 13, Sportivnaya str., Ryazan, 390011, Russia |
| 1700 | SingHealth Centralised Institutional Review Board |  | 150168, Singapore |
| 1800 | Pharma-Ethics Independent Research Ethics Committee |  | Centurion, Gauteng, 0157, South Africa |
| 3700 | CEIM Hospital Doce de Octubre | Avda. Cordoba, s/n | Madrid, 28041, Spain |
| 3701 | CEIM Hospital Doce de Octubre | Avda. Cordoba, s/n | Madrid, 28041, Spain |
| 3702 | CEIM Hospital Doce de Octubre | Avda. Cordoba, s/n | Madrid, 28041, Spain |
| 3703 | CEIM Hospital Doce de Octubre | Avda. Cordoba, s/n | Madrid, 28041, Spain |
| 3704 | CEIM Hospital Doce de Octubre | Avda. Cordoba, s/n | Madrid, 28041, Spain |
| 3705 | CEIM Hospital Doce de Octubre | Avda. Cordoba, s/n | Madrid, 28041, Spain |
| 3706 | CEIM Hospital Doce de Octubre | Avda. Cordoba, s/n | Madrid, 28041, Spain |
| 3707 | CEIM Hospital Doce de Octubre | Avda. Cordoba, s/n | Madrid, 28041, Spain |
| 3708 | CEIM Hospital Doce de Octubre | Avda. Cordoba, s/n | Madrid, 28041, Spain |
| 3709 | CEIM Hospital Doce de Octubre | Avda. Cordoba, s/n | Madrid, 28041, Spain |
| 3710 | CEIM Hospital Doce de Octubre | Avda. Cordoba, s/n | Madrid, 28041, Spain |
| 3711 | CEIM Hospital Doce de Octubre | Avda. Cordoba, s/n | Madrid, 28041, Spain |
| 3800 | Regionala etikprövningsnämnden  i Stockholm | FE289 | Stockholm, 171 77, Sweden |
| 3801 | Regionala etikprövningsnämnden  i Stockholm | FE289 | Stockholm, 171 77, Sweden |
| 3802 | Regionala etikprövningsnämnden  i Stockholm | FE289 | Stockholm, 171 77, Sweden |
| 3803 | Regionala etikprövningsnämnden i Stockholm | FE289 | Stockholm, 171 77, Sweden |
| 3804 | Regionala etikprövningsnämnden i Stockholm | FE289 | Stockholm, 171 77, Sweden |
| 3805 | Regionala etikprövningsnämnden i Stockholm | FE289 | Stockholm, 171 77, Sweden |
| 1861 | Research Ethics Committee | National Taiwan University Hospital | Taipei, 100, Taiwan |
| 1862 | Institutional Review Board | Chang Gung Medical Foundation | Taoyuan, 333, Taiwan |
| 1863 | Institutional Review Board | Taipei Veterans General Hospital | Taipei, 11217, Taiwan |
| 1864 | Institutional Review Board | Taipei Medical University Shuang Ho Hospital | Taipei, 235, Taiwan |
| 1866 | Institutional Review Board | Kaohsiung Medical University Chung-Ho Memorial Hospital | Kaohsiung, 807, Taiwan |
| 1962 | The Institutional Review Board of the Faculty of Medicine, Chulalongkorn University |  | Bangkok, 10330, Thailand |
| 1901 | Baskent University | Clinical Trials Ethical Committee | Ankara, 06490, Turkey |
| 1902 | Baskent University | Clinical Trials Ethical Committee | Ankara, 06490, Turkey |
| 1903 | Baskent University | Clinical Trials Ethical Committee | Ankara, 06490, Turkey |
| 1904 | Baskent University | Clinical Trials Ethical Committee | Ankara, 06490, Turkey |
| 1905 | Baskent University | Clinical Trials Ethical Committee | Ankara, 06490, Turkey |
| 3901 | West of Scotland REC 1 | Clinical Research & Development, West Glasgow Ambulatory Care Hospital | Glasgow, G3 8SJ,  United Kingdom |
| 3902 | West of Scotland REC 1 | Clinical Research & Development, West Glasgow Ambulatory Care Hospital | Glasgow, G3 8SJ,  United Kingdom |
| 5000 | Western Institutional Review Board |  | Olympia, WA, 98502-5010, United States |
| 5001 | Western Institutional Review Board |  | Puyallup, Washington, 98374, United States |
| 5002 | Western Institutional Review Board |  | Puyallup, Washington, 98374, United States |
| 5003 | Avera Institutional Review Board |  | Sioux Falls,South Dakota, 57108, United States |
| 5004 | Indiana University Institutional Review Board |  | Indianapolis, Indiana,  46202-2915, United States |
| 5006 | Edward-Elmhurst Healthcare Institutional Review Board |  | Naperville, Illinois, 60540, United States |
| 5007 | Frederick Memorial Hospital Institutional Review Board |  | Frederick, MD, 21701,  United States |
| 5008 | Western Institutional Review Board |  | Puyallup, Washington, 98374, United States |
| 5010 | Human Subjects Protection office, A115 |  | Hershey, Pennsylvania, 17033-0855, United States |
| 5011 | Cedars-Sinai Medical center; Office of Research Compliance and Quality Improvement |  | Los Angeles, California, 90048, United States |
| 5012 | University of Texas, Southwestern Medical Center Institutional Review Board |  | Dallas, Texas, 75390-8843, United States |
| 5016 | Florida Hospital Institutional Review Board |  | Maitland, Florida, 32751, United States |
| 5017 | Glendale Adventist Medical Center - IRB |  | Glendale, California, 91206, United States |
| 5018 | Western Institutional Review Board |  | Puyallup, Washington, 98374-2115, United States |
| 5019 | Johns Hopkins Institutional Review Board |  | Baltimore, MD, 21205-1911, United States |
| 5022 | US Oncology INC IRB |  | The Woodlands, Texas, 77380, United States |
| 5024 | US Oncology INC IRB |  | The Woodlands, Texas, 77380, United States |
| 5025 | US Oncology INC IRB |  | The Woodlands, Texas, 77380, United States |
| 5026 | US Oncology INC IRB |  | The Woodlands, Texas, 77380, United States |
| 5027 | US Oncology INC IRB |  | The Woodlands, Texas, 77380, United States |
| 5028 | US Oncology INC IRB |  | The Woodlands, Texas, 77380, United States |
| 5029 | US Oncology INC IRB |  | The Woodlands, Texas, 77380, United States |
| 5030 | US Oncology INC IRB |  | The Woodlands, Texas, 77380, United States |
| 5031 | US Oncology INC IRB |  | The Woodlands, Texas, 77380, United States |
| 5032 | US Oncology INC IRB |  | The Woodlands, Texas, 77380, United States |
| 5034 | US Oncology INC IRB |  | The Woodlands, TX, 77380, United States |
| 5036 | US Oncology INC IRB |  | The Woodlands, Texas, 77380, United States |
| 5041 | Western Institutional Review Board |  | Puyallup, Washington,  98374-2115, United States |
| 5042 | WIRB |  | Olympia, WA, 98502-5010, United States |
| 5043 | Western Institutional Review Board |  | Puyallup, Washington, 98374, United States |
| 5044 | Western Institutional Review Board |  | Puyallup, Washington, 98374, United States |
| 5045 | Western Institutional Review Board |  | Olympia, Washington, 98502, United States |
| 5046 | US Oncology INC IRB |  | The Woodlands, Texas, 77380, United States |
| 5056 | Biomedical Research Alliance of New York LLC/Institutional Review Board |  | Lake Success, New York, 11042, United States |
| 5057 | Glens Falls Hospital |  | Glens Falls, New York, 12801, United States |
| 5060 | Ingalls Memorial Hospital | IRB | Harvey, IL, 60426,  United States |
| 5063 | Western Institutional Review Board |  | Puyallup, Washington, 94374, United States |
| 5064 | Quorum review, INC. |  | Seattle, Washington, 98101, United States |
| 5067 | University of Chicago Institutional Review Board |  | Chicago, Illinois, 60637, United States |
| 5068 | Western Institutional Review Board |  | Seatlle, Washington, 98101, United States |
| 5069 | Mercy Institutional Review Board |  | Chesterfield, Missouri, 63017, United States |
| 5070 | University of Colorado Health Intitutional Review Board |  | Fort Collins, Colorado, 80525, United States |
| 5071 | Duke University Health System Institutional Review Board |  | Durham, North Carolina, 22705, United States |
| 5072 | NorthShore University HealthSystem | IRB | Evanston, IL, 60201,  United States |
| 5074 | Institutional Review Board,University of California, Davis |  | Sacramento, California, 95817, United States |
| 5076 | Advarra IRB |  | Colombia, Maryland, 21046, United States |
| 5078 | Western Institutional Review Board |  | Puyallup, WA, 98374,  United States |
| 5080 | The Cooper Health System Institutional Review Board |  | Camden, NJ, 08103,  United States |
| 5083 | Western Institutional Review Board |  | Olympia, Washington, 98502-5010, United States |
| 5087 | Dana-Farber Cancer Institutional review Board,Office for Protection of Research Subjects |  | Boston, Massachusetts, 02215, United States |
| 5088 | Dean Institutional Review Board |  | Middleton, Washington, 53562, United States |
| 5090 | Western Institutional Review Board |  | Olympia, Washington, 98502, United States |
| 5091 | Western Institutional Review Board |  | Puyallup, Washington,  98374-2115, United States |
| 5092 | Western Institutional Review Board |  | Olympia, 98502, United States |
| 5093 | Western Institutional Review Board |  | Olympia, Washington,  98502-5010, United States |
| 5094 | Sutter Health Institutional Review Board |  | Walnut Creek, California, 94596, United States |
| 5095 | U.T. MD Anderson Cancer Center Institutional Review Board |  | Houston, Texas, 77030, United States |
| 5099 | Western Institutional Review Board |  | Puyallup, Washington, 98374, United States |
| 5600 | Vanderbilt University Institutional Review Board |  | Nashville, Tennessee, 37232, United States |
| 5603 | Western Institutional Review Board |  | Olympia, Washington, 98502,United States |
| 5604 | Western Institutional Review Board |  | Puyallup, Washington,  98374-2115, United States |
| 5607 | Western Institutional Review Board |  | Puyallup, WA, 98374, United States |
| 5608 | Western Institutional Review Board |  | Olympia, Washington, 98502-5010, United States |
| 5609 | Western Institutional Review Board |  | Olympia, WA, 98502-5010, United States |
| 5611 | Lehigh Valley Health network IRB |  | Allentown, PA, 18103,  United States |
| 5612 | Western Institutional Review Board |  | Puyallup, Washington, 98374, United States |
| 5614 | Kaiser Permanente Hawaii Institutional Review Board |  | Honolulu, Hawaii, 96813, United States |
| 5616 | Western Institutional Review Board |  | Puyallup, Washington,  98374-2115, United States |
| 5618 | Western Institutional Review Board |  | Puyallup, Washington, 98374-2115, United States |
| 5622 | University of Miami Institutional Review Board |  | Miami, Florida, 33136,  United States |
| 5625 | Biomedical Research Alliance of New York |  | Lake Success, New York, 11042, United States |
| 5628 | Western Institutional Review Board |  | Olympia, WA, 98502-5010, United States |
| 5630 | New York University School of Medicine Institutional Review Board |  | New York, 10016, United States |
| 5634 | Western Institutional Review Board |  | Puyallup, Washington, 98374, United States |
| 5638 | Sacred Heart Health System Institutional Review Board |  | Pensacola, Florida, 32504, United States |

Table 2. List of Independent Ethics Committees (IEC) or Institutional Review Boards (IRB) by study center with ethical approvals for MONALEESA 3

| **Center No.** | **Ethics Committee or  Institutional Review Board** | **Department / Organization** | **City, State/Province, Postal Code Country** |
| --- | --- | --- | --- |
| 1000 | Bellberry Limited | Bellberry Human Research Ethics Committee | Eastwood, SA, 5063 Australia |
| 1002 | Bellberry Limited | Bellberry Human Research Ethics Committee | Eastwood, SA, 5063 Australia |
| 1003 | Bellberry Limited | Bellberry Human Research Ethics Committee | Eastwood, SA, 5063 Australia |
| 1004 | Flinders Medical Centre HREC | Flinders Medical Centre | Bedford Park, SA, 5042 Australia |
| 1200 | Seoul National University Hospital Institutional Review Board |  | Seoul, 03080, Korea,Republic of |
| 1201 | Yonsei University Health System, Severance Hospital, Institutional review board |  | Seoul, 03722, Korea,Republic of |
| 1202 | Samsung Medical Center, Institutional review board |  | Seoul, 06351, Korea,Republic of |
| 1400 | American University of Beirut Medical Center IRB |  | Beirut, Hamra, 1107 2020 Lebanon |
| 1401 | Hotel Dieu de France Hospital IRB |  | Beirut, Achrafiyeh, 166830 Lebanon |
| 1600 | Domain Specific Review Board | Domain B | Singapore, 169039 Singapore |
| 2000 | Ethik-Kommission | Medizinische Fakultaet der Eberhard-Karls-Universitaet und am Universitaetsklinikum Tuebingen | Tuebingen, 72074 Germany |
| 2001 | Ethik-Kommission des Landes Berlin | Landesamt fuer Gesundheit und Soziales | Berlin, 10707 Germany |
| 2002 | Ethik-Kommission | Landesaerztekammer Thueringen | Jena-Maua, 07751 Germany |
| 2003 | Ethik-Kommission | Landesaerztekammer Baden-Wuerttemberg | Stuttgart, 70597 Germany |
| 2004 | Ethik-Kommission | Universitaet Erlangen-Nuernberg Medizinische Fakultaet | Erlangen, 91504 Germany |
| 2005 | Ethik-Kommission | Landesärztekammer Hessen | Frankfurt am Main, 60314 Germany |
| 2006 | Ethik-Kommission des Landes Berlin | Landesamt fuer Gesundheit und Soziales | Berlin, 10707 Germany |
| 2007 | Ethik-Kommission | Aerztekammer Westfalen-Lippe und der Medizinischen Fakultaet der Westfaelischen Wilhelms-Universitaet Muenster | Muenster, 48147 Germany |
| 2008 | Ethik-Kommission | Landesaerztekammer Baden-Wuerttemberg | Stuttgart, 70597 Germany |
| 2009 | Ethikkommission der Ärztekammer Nordrhein |  | Düsseldorf, 40474 Germany |
| 2010 | Ethik-Kommission | Universitaet zu Luebeck Medizinische Fakultaet | Luebeck, 23538 Germany |
| 2011 | Ethik-Kommission | Aerztekammer Nordrhein | Duesseldorf, 40474 Germany |
| 2014 | Ethik-Kommission | Aerztekammer des Saarlandes | Saarbruecken, 66111 Germany |
| 2015 | Ethikkommission | der Landesärztekammer Bayern | München, 81677 Germany |
| 2016 | Ethik-Kommission | Technische Universitaet Dresden Medizinische Fakultaet | Dresden, 01307 Germany |
| 2018 | Ethik-Kommission | Fakultaet fuer Medizin der TU Muenchen | Muenchen 81675 Germany |
| 2019 | Ethikkommission der Ärztekammer Nordrhein |  | Düsseldorf, 40474 Germany |
| 2020 | Ethik-Kommission | Aerztekammer Niedersachsen | Hannover, 30175 Germany |
| 2021 | Ethik-Kommission | Aerztekammer Hamburg | Hamburg, 22083 Germany |
| 2022 | Ethik-Kommission | Bayerische Landesaerztekammer | Muenchen, 81677 Germany |
| 2025 | Ethik-Kommission | Bayerische Landesaerztekammer | Muenchen, 81677 Germany |
| 2026 | Ethik-Kommission | Bayerische Landesaerztekammer | Muenchen 81677 Germany |
| 2027 | Ethik-Kommission | Aerztekammer Hamburg | Hamburg, 22083 Germany |
| 2028 | Ethik-Kommission | Aerztekammer Niedersachsen | Hannover, 30175 Germany |
| 2029 | Ethik-Kommission | Aerztekammer Nordrhein | Duesseldorf, 40474 Germany |
| 2030 | Ethik-Kommission | Aerztekammer Niedersachsen | Hannover, 30175 Germany |
| 2200 | Intezeti Etikai Bizottsag |  | Budapest, 1134 Hungary |
| 2201 | Regionalis es Intezmenyi Kutatasetikai Bizottsag |  | Debrecen, 4012 Hungary |
| 2202 | Kutatasetikai Bizottsag |  | Szolnok, 5000 Hungary |
| 2204 | Tudomanyos es Intezeti Kutatasetikai Bizottsag |  | Budapest, 1032 Hungary |
| 2205 | Intezeti Etikai Bizottsag |  | Budapest, 1122 Hungary |
| 2401 | NRES Committee East of England - Cambridge South |  | Gonville Place, Cambridge, CB1 1LY United Kingdom |
| 2403 | East of England – Cambridge South Research Ethics Commitee | National Research and Ethics Service | Nottingham, NG1 6FS, United Kingdom |
| 2600 | Etická komise MOU |  | Brno, 656 53 Czech Republic |
| 2601 | Etická komise Nemocnice Na Bulovce |  | Prague, 8 180 81 Czech Republic |
| 2602 | Etická komise Všeobecné fakultní nemocnice v Praze |  | Prague 2, 128 08 Czech Republic |
| 2603 | Etická komise pro multicentrické klinické hodnocení Krajská nemocnice Liberec |  | Liberec, 460 63 Czech Republic |
| 2604 | Etická komise Fakultní nemocnice Brno |  | Brno, 625 00 Czech Republic |
| 2800 | King Hussein Cancer Center Institutional Review Board |  | Amman, 11941 Jordan |
| 3000 | CPP Est IV |  | Strasbourg, 67091 France |
| 3002 | CPP Est IV |  | Strasbourg, 67091 France |
| 3003 | CPP Est IV |  | Strasbourg, 67091 France |
| 3006 | CPP Est IV |  | Strasbourg, 67091 France |
| 3009 | CPP Est IV |  | Strasbourg, 67091 France |
| 3010 | CPP Est IV |  | Strasbourg, 67091 France |
| 3013 | CPP Est IV |  | Strasbourg, 67091 France |
| 3014 | CPP Est IV |  | Strasbourg, 67091 France |
| 3015 | CPP Est IV |  | Strasbourg, 67091 France |
| 3016 | CPP Est IV |  | Strasbourg, 67091 France |
| 3017 | CPP Est IV |  | Strasbourg, 67091 France |
| 3018 | CPP Est IV |  | Strasbourg, 67091 France |
| 3019 | CPP Est IV |  | Strasbourg, 67091 France |
| 3023 | CPP Est IV |  | Strasbourg, 67091 France |
| 3200 | Comité d’éthique hospitalo-facultaire - CHU de Liège |  | Liège, 4000 Belgium |
| 3201 | Commissie Medische Ethiek van de UZ Leuven |  | Leuven, 3000 Belgium |
| 3202 | Ethische Toetsingscommissie van het Jessa Ziekenhuis |  | Hasselt, 3500 Belgium |
| 3203 | Comité d’éthique de la CMSE Namur |  | Namur, 5000 Belgium |
| 3204 | Comité d’éthique du GHdC |  | Charleroi,6000 Belgium |
| 3205 | Ethisch Comité van het OLVZ Aalst |  | Aalst, 9300 Belgium |
| 3401 | Ethikkommission der Stadt Wien |  | Vienna, 1030 Austria |
| 3402 | Ethikkommission der Medizinischen Universität Wien |  | Wien, 1090 Austria |
| 3404 | Ethikkommission der medizinischen Universität Innsbruck |  | Innsbruck, 6020 Austria |
| 3600 | Regional komite for medisinsk og helsefaglig forskningsetikk |  | Oslo,484 Norway |
| 3800 | De Videnskabsetiske Komiteer | De Videnskabsetiske Komiteer for Region Syddanmark | Vejle, Region Syd, 7100 Denmark |
| 3801 | De Videnskabsetiske Komiteer | De Videnskabsetiske Komiteer for Region Syddanmark | Vejle, Region Syd, 7100 Denmark |
| 3802 | De Videnskabsetiske Komiteer for Region Syddanmark | De Videnskabsetiske Komiteer for Region Syddanmark | Vejle, Region Syd, 7100 Denmark |
| 3803 | De Videnskabsetiske Komiteer for Region Syddanmark | De Videnskabsetiske Komiteer for Region Syddanmark | Vejle, Region Syd, 7100 Denmark |
| 3804 | De Videnskabsetiske Komiteer for Region Syddanmark | De Videnskabsetiske Komiteer for Region Syddanmark | Vejle, Region Syd, 7100 Denmark |
| 3805 | De Videnskabsetiske Komiteer for Region Syddanmark | De Videnskabsetiske Komiteer for Region Syddanmark | Vejle, Region Syd, 7100 Denmark |
| 4000 | Comitato Etico | Istituto Clinico Humanitas | Rozzano, Milano,20089 Italy |
| 4002 | COMITATO ETICO DELLA PROVINCIA DI BRESCIA | AO Spedali Civili | Brescia, BS, 25123 Italy |
| 4003 | Spettabile Comitato Etico Indipendente | Fondazione IRCCS Istituto Nazionale dei Tumori di Milano | Milano,20133 Italy |
| 4004 | Spettabile Comitato Catania 2 | c/o AORNAS Garibaldi PiazzaS.M | Catania,95125 Italy |
| 4006 | Spettabile Comitato della ASL LE di Lecce |  | Lecce, Le, 73100 Italy |
| 4007 | Comitato Etico | Istituto Nazioonale per lo Studio e la Cura dei Tumori “Fondazione G. Pascale” Via Mariano Semmola | Napoli,80131 Italy |
| 4008 | Spettabile Comitato per la Sperimentazione Clinica dei Medicinali Area Vasta Nord Ovest | Azienda Ospedaliero Universitaria Pisana di Pisa | Pisa,56126 Italy |
| 4010 | Comitato Etico per le Province di L’Aquila e Teramo | ASL1 Avezzano/Sulmona/L’Aquila | Avezzano, AQ,67051 Italy |
| 4200 | Amphia Academy (AMOA) |  | Breda, 4818 CK Netherlands |
| 4201 | PTC |  | Amsterdam,1066 CX Netherlands |
| 4202 | RvB MST |  | Enschede, 7513 JX Netherlands |
| 4203 | MEC Antonius |  | Nieuwegein, 3435 CM Netherlands |
| 4204 | Zuyderland Medisch Centrum, locatie Heerlen |  | Sittard-Geleen 6162 BG Netherlands |
| 4205 | ETZ TweeSteden |  | Tilburg, 5042 AD Netherlands |
| 4206 | METC azM/UM |  | Maastricht, 6229 EV Netherlands |
| 4207 | MEC Groningen |  | Groningen, 9728 NZ Netherlands |
| 4208 | BoD Deventer hospital |  | Deventer, 7416 SE Netherlands |
| 4213 | ACLU |  | Hoofddorp, 2134 TM Netherlands |
| 4216 | Trial begeleidings commissie |  | Roermond, 6043 CV Netherlands |
| 4218 | METC Zuidwest Holland |  | Den haag, 2545 CH Netherlands |
| 4400 | Ethikkommission Nordwest- und Zentralschweiz (EKNZ) |  | Basel, CH-4056 Switzerland |
| 4405 | Kantonale Ethikkommission Zürich |  | Zurich, CH-8090 Switzerland |
| 4406 | Ethikkommission Nordwest- und Zentralschweiz (EKNZ) |  | Basel, CH-4056 Switzerland |
| 4600 | Comité Coordinador de Ética de la Investigación Biomédica de Andalucía | Consejería de Salud | Sevilla, 41020 Spain |
| 4601 | Hospital Univ. Puerta de Hierro Majadahonda | Secretaría Técnica del CEIC | Majadahonda, Madrid, 28222 Spain |
| 4602 | Agencia de Ensayos Clínicos Servicio de Farmacia | Hospital Clínic de Barcelona | Barcelona, 08036 Spain |
| 4603 | CEIC Área 1 Fundación Investigación Biomédica | Hospital General Gregorio Marañon | Madrid, 28007 Spain |
| 4604 | Hospital Universitario La Paz Secretaría Técnica del Comité Ético de Investigación Clínica |  | Madrid, 28046 Spain |
| 4605 | Comité Coordinador de Ética de la Investigación Biomédica de Andalucía | Consejería de Salud | Sevilla, 41020 Spain |
| 4606 | Comité Ético De Investigación Clínica Hospital Universitario De Salamanca |  | Salamanca, 37007 Spain |
| 4609 | Comité Coordinador de Ética de la Investigación Biomédica de Andalucía | Consejería de Salud | Sevilla, 41020 Spain |
| 4610 | Comité Coordinador de Ética de la Investigación Biomédica de Andalucía | Consejería de Salud | Sevilla, 41020 Spain |
| 4612 | Comité Ético de Investigación Clínica | Área 8- Fundación Hospital de Alcorcón | Alcorcon, Madrid, 28922 Spain |
| 4613 | Comité Ético Investigación Clínica De Galicia | Subdirección de Farmacia y Productos Sanitarios Conselleria de Sanidade | Santiago de Compostela, 15703 Spain |
| 4614 | Ceic – Consorci Sanitari Integral | Hospital General de l´Hospitalet | Barcelona,8906 Spain |
| 4800 | Regionala Etikprövningsnämnden i Stockholm | FE 289 | Stockholm,17177 Sweden |
| 4801 | Regionala Etikprövningsnämnden i Stockholm | FE 289 | Stockholm,17177 Sweden |
| 4803 | Regionala Etikprövningsnämnden i Stockholm | FE 289 | Stockholm,17177 Sweden |
| 5000 | Penn State Milton S. Hershey Medical Center, Penn State College of Medicine | Human Subjects Protection Office | Hershey, PA, 17033 United States |
| 5001 | Qurom IRB |  | Seattle, WA, 98101 United States |
| 5002 | Qurom IRB |  | Seattle, WA, 98101 United States |
| 5004 | Qurom IRB |  | Seattle, WA, 98101 United States |
| 5005 | Quorum Review, Inc. |  | Seattle WA 98101 United States |
| 5007 | NYU School of Medicine IRB |  | New York, NY, 10016 United States |
| 5009 | Quorum Review IRB |  | Seattle, WA, 98101 United States |
| 5010 | Qurom IRB |  | Seattle WA 98101 United States |
| 5011 | Western Institutional Review Board |  | Puyallup WA 98374 United States |
| 5012 | Kaiser Permanente Hawaii Institutional Review Board |  | Honolulu, HI,96817 United States |
| 5013 | Quorum Review, Inc. |  | Seattle, WA, 98101 United States |
| 5014 | Quorum Review IRB |  | Seattle, WA, 98101 United States |
| 5016 | Qurom IRB |  | Seattle, WA, 98101 United States |
| 5020 | UCLA Office of the Human Research Protection Program (OHRPP) |  | Los Angeles, CA, 90095 United States |
| 5026 | Glen Falls Hospital |  | Glen Falls, New York, 12801 United States |
| 5027 | Quorum |  | Seattle, WA, 98101 United States |
| 5029 | Genesis Healthcare System IRB |  | Zanesville, Ohio, 43701 United States |
| 5031 | Western Institutional Review Board |  | Puyallup, WA, 983742115 United States |
| 5032 | University of Colorado Health Institutional Review Board |  | Fort Collin, CO,80525 United States |
| 5033 | Western Institutional Review Board |  | Puyallup, WA, 983742115 United States |
| 5034 | Quorum |  | Seattle, WA, 98101 United States |
| 5038 | Quorum Review Inc. (QIRB) |  | Seattle, WA, 98101 United States |
| 5041 | Western Institutional Review Board |  | Puyallup, WA, 983742115 United States |
| 5042 | Western Institutional Review Board |  | Puyallup WA 983742115 United States |
| 5046 | Quorum Review IRB |  | Seattle, WA, 98101 United States |
| 5051 | Quorum Review IRB |  | Seattle, WA, 98101 United States |
| 6000 | IRB Services |  | 372 Hollandandview suite 300 Aurora, Ontario, L4G 0A5 Canada |
| 6001 | Nova Scotia Health Authority Research Ethics Board | Centre for Clinical Research | Halifax, NS, B3H 1V7 Canada |
| 6002 | Research Ethics Board of the Vitalité Health Network |  | Bathurst, New-Brunswick, E2A 1A9 Canada |
| 6005 | OCREB |  | Toronto, ON, M5G0A3 Canada |
| 6004 | OCREB |  | Toronto, ON, M5G0A3 Canada |
| 6006 | Ontario Cancer Research Ethics Board |  | Toronto, ON, MSG0A3 Canada |
| 6007 | UBC BCCA Research Ethics Board (BCCA REB) Fairmont Medical Building | 750 West Broadway,Suite 1315 | Vancouver, British Columbia, V5Z 1H5 Canada |
| 6008 | UBC BCCA Research Ethics Board (BCCA REB) Fairmont Medical Building | 750 West Broadway, Suite 1315 | Vancouver, British Columbia, V5Z 1H5 Canada |
| 6009 | UBC BCCA Research Ethics Board | 750 West Broadway, Suite 902 | Vancouver, BC V5Z 1H8 Canada |
| 6010 | Comité d'éthique de la recherche du Centre de Santé et de Services Sociaux de Trois Rivières | Main centre, 1991 Boul du Carmel | Trois-Rivières, Quebec, G8Z 1V2 Canada |
| 6011 | Comité d'éthique de la recherche du Centre de Santé et de Services Sociaux de Trois-Rivières | Main centre, 1991 Boul du Carmel | Trois-Rivières, Quebec, G8Z 1V2 Canada |
| 6012 | Comité d’Éthique de la Recherche du CIUSSS de l’Estrie – CHUS | Main Centre, 3001, 12th avenue North, office 2929 | Sherbrooke, QC, J1H 5N4 Canada |
| 6600 | Comité de Etica e Investigaciones |  | Monteria, Cordoba 2300, Colombia |
| 6601 | Comité de ética en Investigación Clínica |  | Bogotá DC,110221 Colombia |
| 6802 | Oaxaca Site Management Organization, S.C. | Oaxaca Site Management Organization, S.C. | Oaxaca, 6800 Mexico |
| 8201 | Ethics Committee for Multicenter Trials |  | Sofia,1000 Bulgaria |
| 8202 | Ethics Committee for Multicenter Trials |  | Sofia,1000 Bulgaria |
| 8204 | Ethics Committee for Multicenter Trials |  | Sofia, 1000 Bulgaria |
| 8600 | Medical Research and Ethics Committee |  | Shah Alam,40170 Malaysia |
| 8601 | Medical Research and Ethics Committee |  | Shah Alam, 40170 Malaysia |
| 8801 | Komisja Bioetyczna przy Okregowej Izbie Lekarskiej |  | Krakow, 31-123 Poland |
| 8804 | Komisja Bioetyczna przy Okregowej Izbie Lekarskiej |  | Krakow, 31-123 Poland |
| 9000 | Comissão de Ética para a Investigação Clínica (CEIC) |  | Lisbon, 1749-004 Portugal |
| 9001 | Comissão de Ética para a Investigação Clínica (CEIC) |  | Lisbon, 1749-004 Portugal |
| 9200 | Ethic committee under Arkhangelsk clinical oncology dispensary |  | Arkhangelsk,163045 Russia |
| 9204 | Independent Ethic Committee | Tambov regional clinical oncology dispensary | Tambov,392000 Russia |
| 9400 | Institutional Review Board | Faculty of Medicine, Chulalongkorn University | Bangkok, 10330 Thailand |
| 9401 | Central Research Ethics Committee |  | Bangkok, 10900 Thailand |
| 9600 | Istanbul University Cerrahpaşa Medical Faculty Clinical Research Ethics Committee | Clinical Research | Istanbul, NA,34098 Turkey |
| 9602 | Istanbul University Cerrahpaşa Medical Faculty Clinical Research Ethics Committee | Clinical Research | Istanbul, NA, 34098 Turkey |
| 9603 | Istanbul University Cerrahpaşa Medical Faculty Clinical Research Ethics Committee | Clinical Research | Istanbul, NA, 34098 Turkey |
| 9604 | Istanbul University Cerrahpaşa Medical Faculty Clinical Research Ethics Committee | Clinical Research | Istanbul, NA, 34098 Turkey |

Table 3. List of Independent Ethics Committees (IEC) or Institutional Review Boards (IRB) by study center with ethical approvals for MONALEESA 7

| **Center No.** | **Ethics Committee or Institutional Review Board** | **Department / Organization** | **Address Country** |
| --- | --- | --- | --- |
| 1000 | Eastern Health | Research Governance and Ethics | Box Hill VIC 3128 Australia |
| 1001 | Calvary health care Newcastle Limited | Research Governance and Ethics | Waratah NSW 2298 Australia |
| 1002 | St John of God health Care Human Research Ethics Committee | Human Research Ethics Committee | Wembley WA 6014 Australia |
| 1003 | Austin health | Human Research Ethics Committee | Heidelberg VIC 3084 Australia |
| 1004 | Bell berry Human Research Ethics Committee | Human Research Ethics Committee | Eastwood SA 5063 Australia |
| 1200 | Joint Chinese University of Hong Kong-New Territories East Cluster Clinical Research Ethics Committee | NA | Hong Kong NA NA Hong Kong |
| 1201 | Research Ethics Committee (Kowloon Central / Kowloon East) | NA | Hong Kong NA NA Hong Kong |
| 1203 | UNIMED Medical Institute Institutional Review Board | NA | Hong Kong NA NA Hong Kong |
| 1301 | Institutional Ethics Committee | NA | Bangalore Karnataka 560095 India |
| 1302 | Manavta Clinical Research Institute Ethics Committee | NA | Nashik Maharashtra 422001 India |
| 1304 | 1. Institutional Ethics Committee 1, 2. Institutional Ethics Committee 2 | NA | Mumbai Maharashtra  India |
| 1305 | Ethical Committee | NA | Kolkata West Bengal 700053 India |
| 1400 | Institutional Review Board | Yonsei University Health System -Severance Hospital | Seoul NA 03722 Korea, republic of |
| 1401 | Seoul National University Bundang Hospital Institutional Review Board | Clinical Trial Center of Seoul National University Bundang Hospital | Seongnam-Si Gyeonggi-Do  South Korea |
| 1402 | Institutional Review Board | Samsung Medical center | Seoul NA 06351 Korea, Republic of |
| 1403 | Asan Medical Center Institutional Review Board | Clinical Trial Center of Asan Medical Center | Seoul Seoul  South Korea |
| 1404 | Institutional Review Board | National Cancer center | Goyang-si Gyeonggi-do 10408 Korea, Republic of |
| 1405 | Seoul National University Hospital Institutional Review Board | Center for Human Research Protection of Seoul National University Hospital | Seoul Seoul  03080  South Korea |
| 1500 | Hammoud Hospital IRB | Hammoud Hospital | Saida Saida NA Lebanon |
| 1501 | American University of Beirut IRB | American University of Beirut | Beirut Beirut NA Lebanon |
| 1502 | Ethics Committee-Hotel Dieu De France | Saint Joseph University | Beirut Beirut NA Lebanon |
| 1503 | Bellevue Medical Center IRB | Bellevue Medical Center | Beirut Beirut NA Lebanon |
| 1504 | Saint Georges Hospital IRB | University of Balamand | Beirut Beirut NA Lebanon |
| 1505 | Ain Wazein IRB | Ain Wazein Hospital | Al Chouf Al Chouf NA Lebanon |
| 1601 | LEC under Arkhangelsk Clinical Oncology Dispensary | Arkhangelsk Oncology Regional Center 145/1, Obvodniy Kanal, Arkhangelsk, Russia, 163045 | Arkhangelsk NA  Russia |
| 1603 | LEC under BioEq | BioEq, LLC,  23, Krasnogvardeyskiy, per. Saint-Petersburg, Russia, 197342 | Saint-Petersburg NA  Russia |
| 1702 | King Abdulaziz Medical City, KAIMRC, National Guard Health affairs | Biomedical Ethics section | Riyadh Central 22490 Saudi Arabia |
| 1800 | Centralised Institutional Review Board | CIRB B | NA NA  Singapore |
| 1801 | Centralised Institutional Review Board | CIRB B | NA NA  Singapore |
| 1900 | Chulalongkorn Institutional Review Board | Faculty of medicine, Chulalongkorn University | Patumwan Bangkok  Thailand |
| 1901 | Siriraj Institutional Review Board | Faculty of Medicine Siriraj Hospital, Mahidol University | Bangkoknoi Bangkok 10700 Thailand |
| 2000 | Research Ethics Committee,  National Taiwan University Hospital | Research Ethics Committee C | Taipei NA 100 R.O.C |
| 2001 | Taipei Veterans General Hospital Institutional Review Board | Institutional Review Board | Taipei City NA  R.O.C |
| 2002 | Taipei Medical University – Joint Institutional Review Board | NA | New Taipei City NA  R.O.C |
| 2003 | Mackay Memorial Hospital Institutional Review Board | Institutional Review Board (2) | Taipei City NA  R.O.C |
| 2004 | Institutional Review Board Committee  Koo Foundation Sun Yat-Sen Cancer Center | NA | Taipei City NA  R.O.C |
| 2005 | Chang Gung Medical Foundation Institutional Review Board | Institutional Review Board A | Taipei City NA  R.O.C |
| 2006 | Institutional Review Board Committee, Changhua Christian Hospital, Taiwan | Institutional Review Board Committee A | Changhua NA 500 R.O.C |
| 2007 | Chang Gung Medical Foundation Institutional Review Board | Institutional Review Board A | Taipei City NA 10507 R.O.C |
| 2100 | İstanbul Üniversitesi Cerrahpaşa Tıp Fakültesi Klinik Araştırmalar Etik Kurulu | İstanbul Üniversitesi Cerrahpaşa Tıp Fakültesi Dekanlığı | İstanbul Cerrahpaşa 34098 Turkey |
| 2101 | İstanbul Üniversitesi Cerrahpaşa Tıp Fakültesi Klinik Araştırmalar Etik Kurulu | İstanbul Üniversitesi Cerrahpaşa Tıp Fakültesi Dekanlığı | İstanbul Cerrahpaşa 34098 Turkey |
| 2102 | İstanbul Üniversitesi Cerrahpaşa Tıp Fakültesi Klinik Araştırmalar Etik Kurulu | İstanbul Üniversitesi Cerrahpaşa Tıp Fakültesi Dekanlığı | İstanbul Cerrahpaşa 34098 Turkey |
| 2103 | İstanbul Üniversitesi Cerrahpaşa Tıp Fakültesi Klinik Araştırmalar Etik Kurulu | İstanbul Üniversitesi Cerrahpaşa Tıp Fakültesi Dekanlığı | İstanbul Cerrahpaşa 34098 Turkey |
| 2104 | İstanbul Üniversitesi Cerrahpaşa Tıp Fakültesi Klinik Araştırmalar Etik Kurulu | İstanbul Üniversitesi Cerrahpaşa Tıp Fakültesi Dekanlığı | İstanbul Cerrahpaşa 34098 Turkey |
| 2105 | İstanbul Üniversitesi Cerrahpaşa Tıp Fakültesi Klinik Araştırmalar Etik Kurulu | İstanbul Üniversitesi Cerrahpaşa Tıp Fakültesi Dekanlığı | İstanbul Cerrahpaşa 34098 Turkey |
| 2200 | Al Ain Medical District Human Research Ethics Committee | College of medicine UAE University | Al Ain Abu Dhabi NA United Arab Emirates |
| 2500 | Comite de Etica en Investigacion Clinica (CEIC) | NA | Buenos Aires Buenos Aires Larrea 1381 Dept 3 Floor Apt “A” – zip code: C1117ABK Argentina |
| 2501 | Comite Institucional de Etica de la Investigacion en Salud Clin. Reina Fabiola | NA | Cordoba Cordoba Oncativo 1248 – zip code: X5004FHP Argentina |
| 2502 | Comite de Etica en Investigacion Clinica (CEIC) | NA | Buenos Aires Buenos Aires Larrea 1381 Dept 3 Floor Apt “A” – zip code: C1117ABK Argentina |
| 2600 | Comite de Etica en Investigacion Clinica- Clinica del Country | Investigacion Clinica-Clinica del Country | Bogotá Cundinamarca 110221 Colombia |
| 2601 | Comite de Etica e investigaciones de Oncomedica S.A | Investigación Clínica- Oncomedica | Montería Cordoba  230017 Colombia |
| 2700 | Comité de Ética en Investigación de la Clínica Bajío CLINBA, S.C. | Comité de Ética en Investigación de la Clínica Bajío CLINBA, S.C. | Guanajuato Guanajuato 36090 Mexico |
| 2706 | Comité de Ética en Investigación de la Clínica Bajío CLINBA, S.C. | Comité de Ética en Investigación de la Clínica Bajío CLINBA, S.C. | Guanajuato Guanajuato  Mexico |
| 2709 | Comité de ética en Investigación de la Escuela de Medicina del Instituto Tecnológico y de Estudios Superiores de Monterrey | Comité de ética en Investigación de la Escuela de Medicina del Instituto Tecnológico y de Estudios Superiores de Monterrey | Monterrey Nuevo León 64710 Mexico |
| 2800 | Comitê de Ética em Pesquisa da Fundação PIO XII- Hospital de Câncer de Barretos | NA | Barretos SP 14784-400 Brazil |
| 2801 | Hospital A.C. Camargo - Fundação Antonio Prudente Rua Professor Antonio Prudente, 211 – Liberdade | NA | Sao Paulo SP 01509-900 Brazil |
| 2802 | Comitê de Ética em Pesquisa da Universidade de Passo Fundo | Pró reitoria de pesquisa e pós graduação | Passo Fundo Rio Grande do Sul 99052-900 Brazil |
| 2804 | Comitê de Ética em Pesquisa da Universidade Regional do Noroeste do Estado do Rio Grande do Sul -Unijui | NA | Ijui Rio Grande do Sul 98700-000 Brazil |
| 2805 | Comitê de Ética em Pesquisa do Instituto Brasileiro de Controle do Câncer Av. Alcântara Machado, 2576 – Mooca Phone: (11) 3474-4227 | Comitê de Ética em Pesquisa do Instituto Brasileiro de Controle do Câncer Av. Alcântara Machado, 2576 – Mooca Phone: (11) 3474-4227 | São Paulo SP 03102-002 Brazil |
| 2807 | Comitê de Ética em Pesquisa do Centro de Referência da Saúde da Mulher-CRSM | Comitê de Ética em Pesquisa do Centro de Referência da Saúde da Mulher-CRSM | São Paulo SP 01317-010 Brazil |
| 2809 | Comitê de Ética em Pesquisa Envolvendo Seres Humanos- CEP/UEL LABESC - Laboratório Escola - Rodovia Celso Garcia CId, Km 380 (PR445) - Campus Universitário | Comitê de Ética em Pesquisa Envolvendo Seres Humanos- CEP/UEL LABESC - Laboratório Escola - Rodovia Celso Garcia CId, Km 380 (PR445) - Campus Universitário | Londrina PR 86057-970 Brazil |
| 2900 | UBC BCCA Research Ethics Board | Fairmont Medical Building | Vancouver BC V5Z1H8 Canada |
| 2901 | IRB Services 372 Hollandview Trail, Suite 300  Aurora,Ontario | NA | Aurora Ontario L4G 0A5 Canada |
| 2902 | Health Research Ethics Board of Alberta | Cancer Committee | Edmonton AB T5J 4A7 Canada |
| 2903 | OCREB (Ontario Cancer Research Ethics Board) | MaRS Center, 661 University Ave, Suite 510 | Toronto Ontario M5G 0A3 Canada |
| 2904 | Comité d'éthique de la recherche CHU de Québec - Université Laval | NA | Quebec QC G1L 3L5 Canada |
| 2907 | OCREB-Ontario Institute for Cancer Research MaRS Centre 661 University Avenue, Suite 510 | NA | Toronto Ontario M5G 0A3 Canada |
| 3300 | Medical Research Ethics Committee | University of Malaya Medical Centre | Kuala Lumpur Wilayah Persekutuan 59100  Malaysia |
| 3301 | Medical Research & Ethics Committee | Ministry of Health | Kuala Lumpur Wilayah Persekutuan 59000 Malaysia |
| 4000 | Commissie Medische Ethiek | UZ KU Leuven | Leuven NA 3000 Belgium |
| 4001 | Commissie voor Medische Ethiek | GZA Ziekenhuizen | Antwerpen NA 2018 Belgium |
| 4002 | Comité D’ Ethique | Institut Bordet | Bruxelles NA 1000 Belgium |
| 4004 | Comité D’ Ethique | Clinique et maternité Ste. Elisabeth | Namur NA 5000 Belgium |
| 4100 | Central IRB/IEC - Central Institutional Review Board/ Independent Ethics Committee | NA | Sofia Sofia  Bulgaria |
| 4101 | Central IRB/IEC - Central Institutional Review Board/ Independent Ethics Committee | NA | Sofia Sofia  Bulgaria |
| 4104 | Central IRB/IEC - Central Institutional Review Board/ Independent Ethics Committee | NA | Sofia Sofia  Bulgaria |
| 4170 | CPP « Sud-Ouest et Outre-Mer III » - Bordeaux | Groupe Hospitalier Pellegrin Service de Pharmacologie Clinique Bâtiment 1A Place Amélie-Raba-Léon | Bordeaux NA 33076 France |
| 4171 | CPP « Sud-Ouest et Outre-Mer III » - Bordeaux | Groupe Hospitalier Pellegrin Service de Pharmacologie Clinique Bâtiment 1A Place Amélie-Raba-Léon | Bordeaux NA 33076 France |
| 4172 | CPP « Sud-Ouest et Outre-Mer III » - Bordeaux | Groupe Hospitalier Pellegrin Service de Pharmacologie Clinique Bâtiment 1A Place Amélie-Raba-Léon | Bordeaux NA 33076 France |
| 4173 | CPP « Sud-Ouest et Outre-Mer III » - Bordeaux | Groupe Hospitalier Pellegrin Service de Pharmacologie Clinique Bâtiment 1A Place Amélie-Raba-Léon | Bordeaux NA 33076 France |
| 4174 | CPP « Sud-Ouest et Outre-Mer III » - Bordeaux | Groupe Hospitalier Pellegrin Service de Pharmacologie Clinique Bâtiment 1A Place Amélie-Raba-Léon | Bordeaux NA 33076 France |
| 4176 | CPP « Sud-Ouest et Outre-Mer III » - Bordeaux | Groupe Hospitalier Pellegrin Service de Pharmacologie Clinique Bâtiment 1A Place Amélie-Raba-Léon | Bordeaux NA 33076 France |
| 4177 | CPP « Sud-Ouest et Outre-Mer III » - Bordeaux | Groupe Hospitalier Pellegrin Service de Pharmacologie Clinique Bâtiment 1A Place Amélie-Raba-Léon | Bordeaux NA 33076 France |
| 4178 | CPP « Sud-Ouest et Outre-Mer III » - Bordeaux | Groupe Hospitalier Pellegrin Service de Pharmacologie Clinique Bâtiment 1A Place Amélie-Raba-Léon | Bordeaux NA 33076 France |
| 4200 | Ethikkommission bei der Medizinischen Fakultät der LMU München | N/A | München Bayern 80336 Germany |
| 4201 | Ethikkommission an der Medizinischen Fakultät der Universität Leipzig | NA | Leipzig NA 04109 Germany |
| 4203 | Ethik-Kommission der Medizinischen Fakultät der Christian-Albrechts-Universität zu Kiel | NA | Kiel NA 24105 Germany |
| 4204 | Ärztekammer Nordrhein Ethik-Kommission | NA | Düsseldorf NA 40474 Germany |
| 4205 | Ethikkommission der LÄK Baden- Württemberg | NA | Stuttgart NA 70597 Germany |
| 4206 | Ethikkommission an der Technischen Universität Dresden | NA | Dresden NA 01307 Germany |
| 4207 | Ethikkommission der Landesärztekammer Hessen | N/A | Frankfurt Hessen 60488 Germany |
| 4211 | Ethikkommission der Ärztekammer Nordrhein | NA | Düsseldorf NA 40474 Germany |
| 4212 | Ethikkommission der Universität Ulm | N/A | Ulm Baden-Württemberg 89081 Germany |
| 4213 | Ethikkommission der Medizinischen Fakultät der Friedrich-Alexander-Universität Erlangen-Nürnberg | N/A | Erlangen Bayern 91054 Germany |
| 4215 | Ethikkommission der LÄK Thüringen | NA | Jena NA 07751 Germany |
| 4216 | Landesärztekammer Baden-Württemberg Ethik-Kommission | NA | Stuttgart NA 70597 Germany |
| 4300 | Scientific Council of Euromedica General Clinic of Thessaloniki | Scientific Council | Thessaloniki Address: 11 Marias Kallas street 54645 Greece |
| 4301 | Scientific Council of the University General Hospital of Heraklion | Scientific Council | Heraklion Address: Voutes, Stavrakia 711 10 Greece |
| 4400 | Medical Research Council | Ethics committee for Clinical Pharmacology | Budapest NA 1051 Hungary |
| 4401 | Medical Research Council | Ethics committee for Clinical Pharmacology | Budapest NA 1051 Hungary |
| 4402 | Medical Research Council | Ethics committee for Clinical Pharmacology | Budapest NA 1051 Hungary |
| 4403 | Medical Research Council | Ethics committee for Clinical Pharmacology | Budapest NA 1051 Hungary |
| 4405 | Medical Research Council | Ethics committee for Clinical Pharmacology | Budapest NA 1051 Hungary |
| 4406 | Medical Research Council | Ethics committee for Clinical Pharmacology | Budapest NA 1051 Hungary |
| 4500 | Comitato etico centrale dell'irccs fondazione salvatore maugeri (irccs) di Pavia | Irccs fondazione salvatore maugeri (irccs) di pavia | Pavia pavia 27100 Italy |
| 4501 | Comitato etico indipendente dell’azienda ospedaliero-universitaria policlinico s.orsola-malpighi di bologna | NA | Via albertoni pietro, 15-padiglione 3 Bologna Bologna 40138 Italy |
| 4503 | Comitato etico catania 2 c/o azienda ospedaliera di rilievo nazionale e di alta specializzazione garibaldi (aornas garibaldi) | NA | Piazza santa maria gesù, 7 Catania Catania 95125 Italy |
| 4505 | Comitato Etico ASL Lecce | ASL- Lecce | Lecce Lecce 73100 Italy |
| 4506 | Comitato Etico dell’Istituto Nazionale Tumori Fondazione Pascale | Via Mariano Semmola | Napoli NA 80131 Italy |
| 4507 | Comitato Etico Lazio 1 | Az Ospedaliera San Camillo Forlanini – Circonvallazione Gianicolense, 87 | Roma RM 00152 |
| 4509 | Comitato etico unico della provincia di ferrara c/o aou di ferrara | NA | Via aldo moro, 8 Ferrara Ferrara 44124 Italy |
| 4511 | Comitato etico area cremona, mantova e lodi c/o ao di cremona | NA | Via della concordia, 1 Cremona Cremona 26100 Italy |
| 4512 | Comitato Etico Area Vasta Centro | Area Vasta Centro-Toscana | Firenze Firenze 50134 Italy |
| 4513 | Comitato etico regionale della liguria c/o irccs aou san martino-ist istituto nazionale per la ricerca sul cancro | NA | Largo rosanna benzi Genova Genova 16132 Italy |
| 4514 | Comitato Etico IRCCS Candiolo SP142 Km 3.95  10060 Candiolo (TO) | NA | Candiolo - torino to 10060 Italy |
| 4517 | Comitato etico regionale (cer) delle marche c/o azienda ospedaliera universitaria ospedali riuniti umberto i-lancisi-gm salesi di ancona | NA | Via conca Ancona Ancona 60126 Italy |
| 4518 | CEAS Umbria | Umbria | Ellera di Corciano Perugia 06070 Italy |
| 4519 | Comitato etico irst irccs avr | NA | Via pieromaroncelli, 40 meldola forli-cesena 47014 Italy |
| 4522 | Comitato etico per le province di l’aquila e teramo. presidio ospedaliero l’aquila, loc.vetoio, coppito | NA | L’aquila l’aquila 67100 Italy |
| 4523 | Comitato etico degli irccs istituto europeo di oncologia e centro cardiologico monzino | NA | Via giuseppe ripamonti, 435 Milano Milano 20141 Italy |
| 4524 | Comitato etico regionale unico (ceru) c/o azienda ospedaliero-universitaria s.maria della misericordia di udine | NA | Piazzale s.maria della misericordia Udine Udine 33100 Italy |
| 4525 | Comitato Etico AreaVasta Nord Ovest | AreaVasta Nord Ovest- Toscana | Pisa Pisa 56126 Italy |
| 4526 | Comitato Etico università Federico II di Napoli | Via Sergio Pansini, 5 | Napoli NA 80131 Italy |
| 4527 | Comitato Etico fondazione Policlinico Universitario Gemelli | Policlinico Gemelli- Roma | Roma Roma 00168 Italy |
| 4528 | Comitato etico delle azienda sanitarie dell’umbria-segreteria scientifico-amministrativa del ceas umbria | NA | Via della rivoluzione, 16 – ellera di Corciano Corciano Perugia 06073 Italy |
| 4601 | Niezależna Komisja Bioetyczna ds. Badań Naukowych przy Gdańskim Uniwersytecie Medycznym | NA | Gdańsk NA 80-210 Poland |
| 4602 | Niezależna Komisja Bioetyczna ds. Badań Naukowych przy Gdańskim Uniwersytecie Medycznym | NA | Gdańsk NA 80-210 Poland |
| 4700 | Comissão de Ética para a Investigação Clínica (CEIC) | N/A | Lisbon Lisbon 1749-004  Portugal |
| 4701 | Comissão de Ética para a Investigação Clínica (CEIC) | N/A | Lisbon Lisbon 1749-004  Portugal |
| 4702 | Comissão de Ética para a Investigação Clínica (CEIC) | N/A | Lisbon Lisbon 1749-004  Portugal |
| 4703 | Comissão de Ética para a Investigação Clínica (CEIC) | N/A | Lisbon Lisbon 1749-004  Portugal |
| 4704 | Comissão de Ética para a Investigação Clínica (CEIC) | N/A | Lisbon Lisbon 1749-004  Portugal |
| 4900 | Commission Cantonale d’éthique et de la Recherche (CCER) | Hôpitaux Universitaires de Genève | Genève GE 1207 Switzerland |
| 5000 | Washington University Human Research Protection Office | NA | Saint Louis MO 63110 United States |
| 5002 | Office of the Human Research Protection Program (OHRPP) | NA | Los Angeles CA 90095 US |
| 5004 | MD Anderson Cancer Center Institutional Review Board | MD Anderson Cancer Center IRB | Houston Houston 77030 United States |
| 5005 | Quorum | NA | Seattle WA 98101 United States |
| 5006 | Quorum Review IRB | NA | Seattle WA 98101 United States |
| 5010 | Western Institutional Review Board | NA | 1019 39th Avenue SE Suite 120 \| Puyallup WA 98374--2115 United States |
| 5011 | Western Institutional Review Board | NA | Puyallup WA 98374 USA |
| 5013 | The Center For Cancer and Blood Disorders IRB | NA | Fort Worth Texas  USA |
| 5015 | Western Institutional Review Board | NA | 1019 39th Avenue SE Suite 120 \| Puyallup WA 98374--2115 United States |
| 5022 | Quorum Review IRB | NA | Seattle WA 98101 USA |
| 5024 | Quorum Review INC | NA | Seattle WA 98101 USA |
| 5025 | University of Miami Institutional review board | NA | Miami FL 33136 USA |
| 5027 | Dana-Farber Cancer Institute IRB | NA | Valencia CA 91355 United States |
| 5028 | IRBMED | NA | Bldg 520 Room 3214,2800 Plymouth Rd Ann Arbor Ml 48109 USA |
| 5030 | Memorial Healthcare System Institutional Review Board | NA | Hollywood FL 33021 United States |
| 5033 | Western Institutional Review Board | NA | 1019 39th Avenue SE Suite 120 Puyallup WA 98374-2215 USA |
| 5034 | BSD IRB, The University of Chicago Biomedical Sciences Division/University of Chicago Medical Centre | 5751 S. Woodlawn Ave,  McGiffert House, 2nd Floor | Chicago IL 60637 USA |
| 5040 | Quorum IRB | NA | Seattle WA 98101 USA |
| 5043 | Kaiser Permanente Hawaii Institutional Review Board | NA | 711 Kapiolani Blvd., Suite 110, Honolulu HI 96813 USA |
| 5044 | BAMC Institutional Review Board | NA | 3551 Roger Brook Drive, Ft Sam Houston, TX 78234 TX 78234 USA |
| 5045 | Quorum Review IRB | NA | Seattle WA 98101 USA |
| 5050 | Western Institutional Review Board (WIRB) | 1019 39th Avenue SE, Suite 120 | Puyallup WA 98374-2115 USA |
| 5061 | Johns Hopkins Medicine Institutional Review Board | NA | Baltimore MD 21205-1911 USA |
| 5062 | Quorum Review IRB | NA | Seattle WA 98101 USA |
| 5063 | Quorum Review IRB | NA | Seattle WA 98101 USA |
| 5065 | Houston Methodist Research Institutional Review Board | NA | Houston TX 77030 USA |
| 5066 | Penn State Milton S. Hershey Medical Center, Penn State College of Medicine | Human Subject Pretection Office | Hershey PA 17033 USA |
| 5067 | Western Institutional Review Board | NA | Puyallup WA 98374 USA |
| 5068 | Western Institutional Review Board | NA | 1019 39th Ave SE, Puyallup WA 98374 USA |
| 5073 | Duke University Health System IRB | Duke University Medical Center, St. 405, Box 2712 | Durham NC 27705 USA |
| 5076 | San Antonio Institutional Review Board | The University of Texas Health Science Center | San Antonio TX 78229-3900 USA |
| 5078 | Western Institutional Review board | NA | Puyallup WA 98374-2115 USA |
| 5084 | SCL Health-Front Range IRB | NA | 1375 E. 19th Ave, Denver CO 80218 USA |
| 5095 | Western Institutional Review Board | NA | Puyallup WA 98374 USA |
| 5096 | WIRB | 1019 39th Ave SE Suite 120 | Puyallup WA 98374 USA |
| 6000 | CEIC Hospital Clínico San Carlos Doctor Martín Lagos, s/n. Puerta G - 4ª Norte | Hospital Clínico San Carlos Doctor Martín Lagos, s/n. Puerta G - 4ª Norte | Madrid Madrid  Spain |
| 6001 | CEIC Hospital Clínico San Carlos Doctor Martín Lagos, s/n. Puerta G - 4ª Norte | Hospital Clínico San Carlos Doctor Martín Lagos, s/n. Puerta G - 4ª Norte | Madrid Madrid  Spain |
| 6002 | CEIC Hospital Clínico San Carlos Doctor Martín Lagos, s/n. Puerta G - 4ª Norte | Hospital Clínico San Carlos Doctor Martín Lagos, s/n. Puerta G - 4ª Norte | Madrid Madrid  Spain |
| 6003 | CEIC Hospital Clínico San Carlos Doctor Martín Lagos, s/n. Puerta G - 4ª Norte | Hospital Clínico San Carlos Doctor Martín Lagos, s/n. Puerta G - 4ª Norte | Madrid Madrid  Spain |
| 6004 | CEIC Hospital Clínico San Carlos Doctor Martín Lagos, s/n. Puerta G - 4ª Norte | Hospital Clínico San Carlos Doctor Martín Lagos, s/n. Puerta G - 4ª Norte | Madrid Madrid  Spain |
| 6005 | CEIC Hospital Clínico San Carlos Doctor Martín Lagos, s/n. Puerta G - 4ª Norte | Hospital Clínico San Carlos Doctor Martín Lagos, s/n. Puerta G - 4ª Norte | Madrid Madrid  Spain |
| 6006 | CEIC Hospital Clínico San Carlos Doctor Martín Lagos, s/n. Puerta G - 4ª Norte | Hospital Clínico San Carlos Doctor Martín Lagos, s/n. Puerta G - 4ª Norte | Madrid Madrid  Spain |
| 6007 | CEIC Hospital Clínico San Carlos Doctor Martín Lagos, s/n. Puerta G - 4ª Norte | Hospital Clínico San Carlos Doctor Martín Lagos, s/n. Puerta G - 4ª Norte | Madrid Madrid  Spain |
| 6008 | CEIC Hospital Clínico San Carlos Doctor Martín Lagos, s/n. Puerta G - 4ª Norte | Hospital Clínico San Carlos Doctor Martín Lagos, s/n. Puerta G - 4ª Norte | Madrid Madrid  Spain |
| 6009 | CEIC Hospital Clínico San Carlos Doctor Martín Lagos, s/n. Puerta G - 4ª Norte | Hospital Clínico San Carlos Doctor Martín Lagos, s/n. Puerta G - 4ª Norte | Madrid Madrid  Spain |
| 6010 | CEIC Hospital Clínico San Carlos Doctor Martín Lagos, s/n. Puerta G - 4ª Norte | Hospital Clínico San Carlos Doctor Martín Lagos, s/n. Puerta G - 4ª Norte | Madrid Madrid  Spain |
| 6011 | CEIC Hospital Clínico San Carlos Doctor Martín Lagos, s/n. Puerta G - 4ª Norte | Hospital Clínico San Carlos Doctor Martín Lagos, s/n. Puerta G - 4ª Norte | Madrid Madrid  Spain |
| 6013 | CEIC Hospital Clínico San Carlos Doctor Martín Lagos, s/n. Puerta G - 4ª Norte | Hospital Clínico San Carlos Doctor Martín Lagos, s/n. Puerta G - 4ª Norte | Madrid Madrid  Spain |
| 6014 | CEIC Hospital Clínico San Carlos Doctor Martín Lagos, s/n. Puerta G - 4ª Norte | Hospital Clínico San Carlos Doctor Martín Lagos, s/n. Puerta G - 4ª Norte | Madrid Madrid  Spain |
| 6015 | CEIC Hospital Clínico San Carlos Doctor Martín Lagos, s/n. Puerta G - 4ª Norte | Hospital Clínico San Carlos Doctor Martín Lagos, s/n. Puerta G - 4ª Norte | Madrid Madrid  Spain |
| 6016 | CEIC Hospital Clínico San Carlos Doctor Martín Lagos, s/n. Puerta G - 4ª Norte | Hospital Clínico San Carlos Doctor Martín Lagos, s/n. Puerta G - 4ª Norte | Madrid Madrid  Spain |
| 6019 | CEIC Hospital Clínico San Carlos Doctor Martín Lagos, s/n. Puerta G - 4ª Norte | Hospital Clínico San Carlos Doctor Martín Lagos, s/n. Puerta G - 4ª Norte | Madrid Madrid  Spain |
